# A network meta-analysis of randomised controlled trials of antipsychotic medications to assess their comparative efficacy and tolerability in autistic people

**DOI:** 10.64898/2026.05.11.26352928

**Authors:** Shoumitro Deb, Bharati Limbu, José Antonio López López, Meera Roy, Meena Murugan, Ashok Roy, Basma Akrout Brizard, Jacopo Santambrogio

## Abstract

**Background:** A high proportion of autistic people receive off-license antipsychotic medication, often in the absence of a mental illness, primarily for behaviours that challenge, which is a public health concern. Although meta-analyses have been published recently, there is a lack of a comprehensive network meta-analysis to inform clinicians about the relative efficacy and safety of antipsychotic medications.

**Aims:** To conduct a network meta-analysis of available RCTs of antipsychotic medications involving autistic participants to assess the relative efficacy of different antipsychotics and their adverse effects.

**Method:** We searched seven databases and hand-searched ten relevant journals. Two authors independently screened titles, abstracts, and full papers, extracted data, and assessed their quality.

**Results:** We analysed data from 22 RCTs involving 1562 autistic people. The largest mean difference with 95% confidence interval in the Aberrant Behaviour Checklist-Irritability (ABC-I) score compared with placebo was from the combined intervention with risperidone and parent training: -11.16 (-15.13, -7.18), followed by risperidone: -7.59 (-9.22, -5.95), and aripiprazole: -5.59 (−7.18, −4). The largest effect on Clinical Global Impression-Improvement (CGI-I) scores was from risperidone, 7.65 (2.17, 27.04), followed by aripiprazole, 7.02 (1.92, 25.72), compared with placebo. Risperidone (4; 1.57, 10.21) and aripiprazole (2.77; 1.20, 6.39) had significantly higher odds ratios for adverse effects, but aripiprazole showed the least weight gain.

**Conclusions:** Combined parent training and risperidone followed by risperidone and aripiprazole showed the best effects on the ABC-I score, whereas risperidone and aripiprazole showed the greatest effect on the CGI-I score. However, risperidone and aripiprazole showed significantly increased adverse effects.

## Introduction

Autism Spectrum Disorder (ASD) is a neurodevelopmental disorder characterised by impaired communication and repetitive and restrictive behaviour that affects 1 in 45 children and often continues into adulthood (Bertelli et al., 2022). ASD is often comorbid with other neurodevelopmental disorders like intellectual disabilities (ID), attention deficit hyperactivity disorders (ADHD), and psychiatric disorders like psychosis and anxiety disorders. A high proportion of autistic people receive psychotropic medicine for psychiatric disorders as well as behaviours that challenge (BtC), which is common in ASD (Deb, 2024). Psychostimulants are commonly prescribed because of comorbid ADHD, but antipsychotic prescriptions are also prevalent, particularly used for BtC. National and international guidelines recommend the use of non-pharmacological psychological and behavioural interventions such as positive behaviour support (PBS) for BtC before considering psychotropics, including antipsychotics (National Institute for Health and Care Excellence [NICE], 2015; Deb et al., 2009). Therefore, if antipsychotics are used for BtC, which tends to be common, it is important to explore the evidence of their effectiveness and adverse effects profile to help clinicians decide when to use psychotropics, including antipsychotics and also which antipsychotics, if they are needed. We, therefore, conducted a network meta-analysis (NMA) of all published RCTs on antipsychotics for autistic people to assess their relative efficacy and tolerability, as this has not been done before

## Methods

### Search strategy and selection criteria

The protocol for the current NMA has been registered at PROSPERO (crd.york.ac.uk/prospero) (registration number CRD42022343669). For reporting, we followed the Preferred Reporting Items for Systematic Reviews and Meta-analysis Protocol (PRISMA-NMA; see supplementary material 1) and the extension statements for NMA (Hutton et al., 2015). We searched Embase, Medline, Cochrane, Clinicaltrials.gov, PsycINFO, CINAHL, and ERIC databases in English from their inception until 30th May 2022 and updated the search from 30^th^ May 2022 to 1^st^ September 2024. Additionally, we hand-searched four relevant journals in the field of ASD (Autism, Journal of Autism and Developmental Disorders, Autism Research, Research in Autism Spectrum Disorder) and six in psychopharmacology (Psychopharmacology, Neuropsychopharmacology, International Journal of Neuropsychopharmacology, Journal of Clinical Psychopharmacology, Human Psychopharmacology, Journal of Child and Adolescent Psychopharmacology) for relevant articles between 2000 and 1^st^ September 2024. We also searched the bibliographies of identified articles for cross-referencing. We searched for grey literature, including conference abstracts and unpublished data on clinicaltrials.gov site. The search strategy and search terms are presented in supplementary material 1.

We included randomised controlled trials (RCTs) of any antipsychotics involving autistic people (defined using a standardised method) of all ages for any outcome (e.g. psychiatric disorder, behaviours that challenge and ASD core symptoms). The control intervention could be a placebo, another medication or a non-pharmacological intervention. Crossover trials were included only if data were available from Phase I to avoid carryover effects in Phase II. We contacted the authors for missing data.

### Data synthesis

Using the eligibility criteria, two authors (MR and JS) independently screened titles, abstracts and full papers (supplementary material 1). Using the Cochrane Risk of Bias 2.0 (RoB) Scale, two authors (AR and MM) independently assessed the quality of papers (Sterne et al., 2019). Using a data extraction form based on the Cochrane Handbook template (Li et al., 2024), two authors (BAB and MM) independently extracted data from the selected papers (supplementary material 1). Initial data extraction started on 21^st^ July 2022. A third author (SD) arbitrated any discrepancies between the two reviewers that could not be resolved through discussion.

### Outcomes

#### Primary outcome

Primary outcomes included (a) any psychopathology, including psychiatric disorders such as psychoses, schizophrenia, etc., (b) ASD core symptoms such as impairment in social and other communications and restrictive and repetitive behaviours and (c) associated behaviours such as agitation, irritability and aggression.

#### Secondary outcomes

Secondary outcomes included (a) behaviours such as sleep disturbance, self-mutilation, attention and concentration problems, (b) global assessment of health and function, (c) QoL (quality of life) of the participants and their families, (d) adverse effects, (e) dropout rate due to any cause, and (f) dropout rate due to adverse effects.

### Statistical methods and analysis

We conducted the NMA within a frequentist framework using MetaInsight: The Complex Review Support Unit (CRSU) network meta-analysis (NMA) web-based app (Owen et al., 2019). We conducted a descriptive assessment of the transitivity assumption by carefully exploring and examining the characteristics, design, and covariates of each included study in the network (supplementary material 2). Covariates included the medication being a novel treatment for the individual, the presence and severity of ID, age, presence of psychiatric disorders and BtC. Inconsistency and heterogeneity were also assessed in MetaInsight CRSU. We conducted sensitivity analyses for high heterogeneity as pre-specified in the protocol. We performed fixed and random-effects models for the NMA and reported findings from the more conservative model. We ranked interventions using P-scores from the ‘netmeta’ package in R (Rücker & Schwarzer, 2015; Balduzzi et al., 2023; Shim et al., 2019), with values ranging from 0 to 1, where higher values indicate greater intervention efficacy. We used intention-to-treat analysis results (where available) for continuous and dichotomous outcomes. For continuous outcomes, we used the mean and standard deviation (SD) change from baseline to follow-up, and the endpoint mean and SD for studies that didn’t report baseline-to-follow-up change. We combined the data using the mean difference (MD) as per the Cochrane Handbook guidelines (Deeks et al., 2024). We decided to use the combination method to maximise the number of studies included, as no appropriate conversion method was available. We did not exclude studies due to incomplete data, as this may introduce bias in the NMA results.

We calculated MD to summarise intervention effects for continuous outcomes, with 95% CIs (Confidence Intervals). We calculated odds ratios (ORs) and 95% CIs for dichotomous outcomes. We did not combine outcomes that reported continuous and dichotomous data. Therefore, we analysed the data type with the most available data points. We excluded studies with insufficient data and those for which we could not convert data. All multi-arm studies with different doses were combined to form a single node for each medication type. The outcome data from multi-arm studies were pooled to produce two-arm interventions. Only two-arm studies were included in the NMA to improve network structure and connectivity. Separate NMAs were conducted based on different primary and secondary outcomes to ensure transitivity. We assessed each study’s risk of bias (RoB) using the Cochrane RoB 2.0 tool (Sterne et al., 2019) and conducted a sensitivity analysis, excluding studies with a high or unclear RoB. A narrative summary of the findings is presented in supplementary material 3.

## Results

### Study selection and characteristics of included studies

We identified 2340 citations from the initial search, of which 523 were duplicates. In the end, we included 39 studies in the review, of which 21 were primary and 18 were secondary, originating from the 21 primary RCTs (Figure 1) (Deb et al., 2023). We identified 622 more citations from our updated search, of which only one was eventually included (Figure 1). In total, 22 RCTS were included in the current NMA (Aman et al., 2009; DeVane et al., 2019; Ghanizadeh et al., 2013; Hollander et al., 2006; Ichikawaet al., 2016; Kent et al., 2013; Kouhbanani et al., 2021; Loebel et al., 2016; Luby et al., 2006; Mahajan et al., 2022; Marcus et al., 2009; Martsenkovska & Martsenkovska, 2014; McCracken et al., 2002; McDougle et al., 1998; Miral et al., 2008; Nagaraj et al., 2006; NCT00198107; NCT00468130; NCT01624675; Nikvarz et al., 2016; Owen et al., 2009; Shea et al., 2004). The reasons for excluding studies from the NMA are provided in supplementary material 1. Data from 1562 autistic people were included in these primary studies, of whom 1326 completed the study (85%). Only primary studies were included in the NMA. The characteristics of the primary RCTs are reported in the narrative summary (supplementary material 1). The characteristics and results of the secondary articles are reported in a previous publication (Deb et al., 2023).

**Figure 1:**
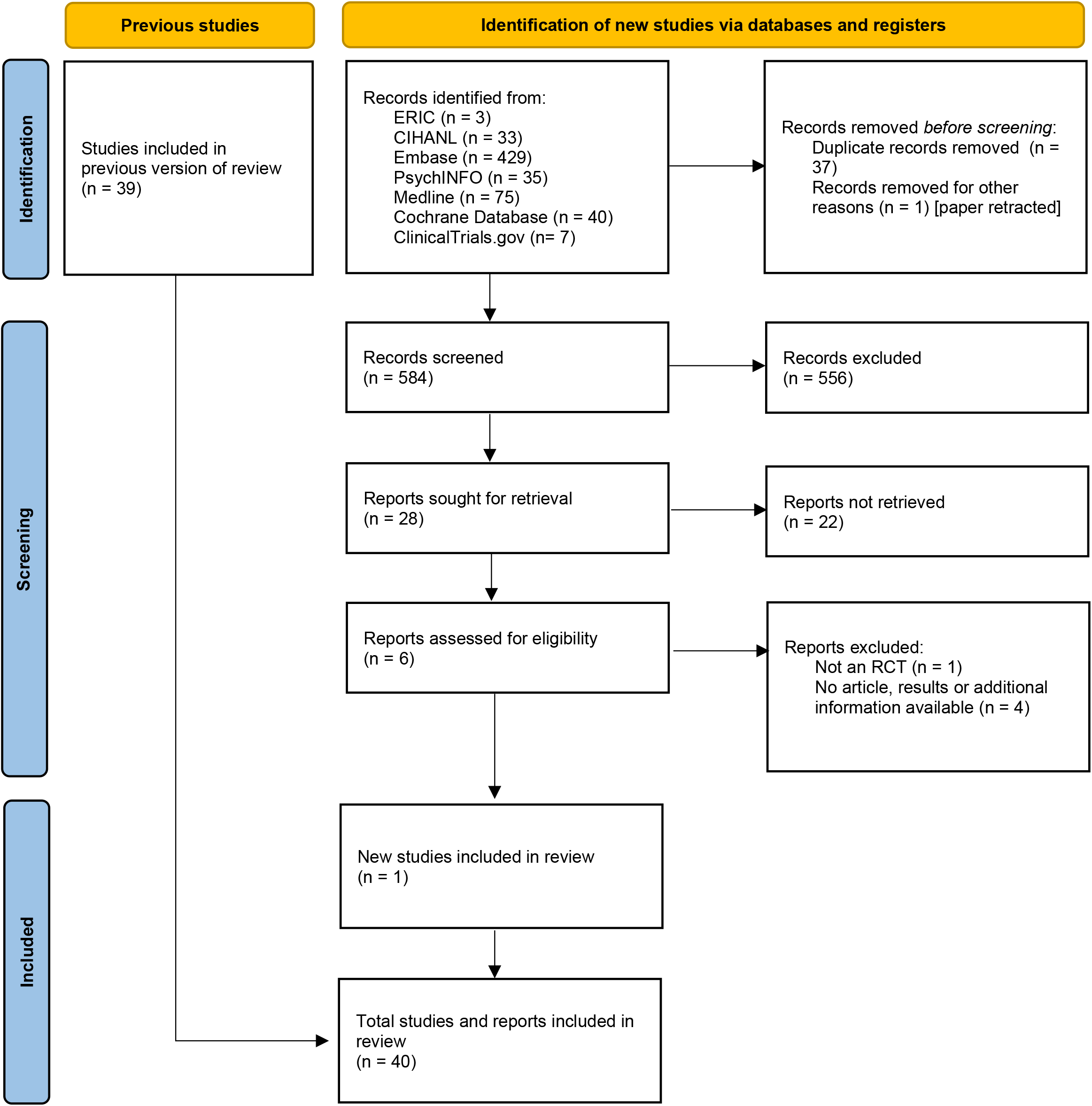
Study selection.

Apart from one small study (n=31) (McDougle et al., 1998) that included adults aged 18 and older, the rest included only children and adolescents (2-17.5 years). There was wide variation in the sample sizes in the included studies, ranging from 11 to 218, with a median of 60. Most participants were males (81.3%). Data on race were available from 12 studies (Aman et al., 2009; DeVane et al., 2019; Hollander et al., 2006; Kent et al., 2013; Loebel et al., 2016; Luby et al., 2006; Mahajan et al., 2022; Marcus et al., 2009; McCracken et al., 2002; McDougle et al., 1998; Owen et al., 2009; Shea et al., 2004). Across these studies, the majority of participants were White (N=714), followed by Black (N=177), Asian (N=68), and Hispanic (N=25). Additionally, 48 reported their background as other, and 3 reported it as mixed-race. Data on socio-economic status (SES) were available from four studies (Luby et al., 2006; Mahajan et al., 2022; McCracken et al., 2002; Nagaraj et al., 2006), with most participants classified as having a very high SES (N= 94), followed by middle-low (N= 44), middle-high (N=39) and low (N=16). Of the 15 RCTs on risperidone, seven compared risperidone with placebo (Kent et al., 2013; Luby et al., 2006; McCracken, 2002; McDougle et al., 1998; Nagaraj et al., 2006; NCT01624675; Shea et al., 2004), two with aripiprazole (DeVane et al., 2019; Ghanizadeh et al., 2013), one each with haloperidol (Miral et al., 2008), methylphenidate (Mahajan et al., 2022), memantine (Nikvarz et al., 2016), divalproex sodium (Martsenkovska & Martsenkovska, 2014), a combination of risperidone and parent education (Aman et al., 2009), and a combination of risperidone and virtual reality relaxation training (Kouhbanani et al., 2021). There were seven RCTs involving aripiprazole and one each involving olanzapine (Hollander et al., 2006) and lurasidone (Loebel et al., 2016). Five of the seven RCTs compared aripiprazole with a placebo (Ichikawa et al., 2016; Marcus et al., 2009; NCT00198107; NCT00468130; Owen et al., 2009), and the other two compared aripiprazole with risperidone (DeVane et al., 2019; Ghanizadeh et al., 2013). Most studies did not report participants’ IQ, and only nine included individuals with ID. The Aberrant Behavior Checklist-Irritability subscale (ABC-I) (Aman et al., 1995) and Clinical Global Intervention-Improvement (CGI-I) scale (National Institute of Mental Health, 1976) were the most commonly used outcome measures in the majority of the studies to assess the effect of antipsychotics on associated behaviours such as irritability, aggression and agitation.

Eight studies assessed (Aman et al., 2009; Hollander et al., 2006; Ichikawa et al., 2016; Kent et al., 2013; Loebel et al., 2016; Marcus et al., 2009; NCT00198107; Owen et al., 2009) compulsive behaviour using the Children-Yale Brown Obsessive Compulsive Scale (C-YBOCS) (Scahill et al., 1997), and one study (McDougle et al., 1998) assessed compulsive behaviour using the Yale-Brown Obsessive Compulsive Scale (YBOCS) (Goodman et al., 1989). We did not report the C-YBOCS data for NMA because, in most studies, this scale was not used to measure obsessive-compulsive symptoms but was used as a proxy measure for ASD core symptoms, such as restrictive and repetitive behaviour. Results are provided in supplementary material 3.

Only three studies assessed the effect of antipsychotic medications on social functioning using the Social Reciprocity Scale (Constantino et al., 2003) (NCT00198107) and the Ritvo-Freeman Real Life Rating Scale (Freeman et al., 1986) (McDougle et al., 1998; Miral et al., 2008). However, there was not enough data to conduct an NMA. No RCT assessed the effect of medications on psychiatric disorders such as psychoses or bipolar disorder. The RoB rating is presented in supplementary material 1. Many studies (n=9) were judged to have some concerns regarding the overall RoB. Eight received a high (Kouhbanani et al., 2021; Luby et al., 2006; Mahajan et al., 2022; Martsenkovska & Martsenkovska, 2014; Miral et al., 2008; NCT00198107; NCT00468130; Nikvarz et al., 2016) and five a low overall risk (Ichikawa et al., 2016; Kent et al., 2013; Loebel et al., 2016; Nagaraj et al., 2006; Owen et al., 2009). There was no evidence of inconsistency in any of the NMA analyses conducted where both direct and indirect comparisons were available (supplementary material 3). However, inconsistency could not be evaluated for comparisons based only on indirect evidence. Based on our descriptive assessment of transitivity, we did not detect any issues with the majority of the covariates. However, we did not find sufficient information on the presence of ID in the primary studies. Pairwise meta-analyses have also been reported in supplementary material 3.

### Primary outcomes

Figure 2 presents the network structure, forest plots, and NMA results for the clinical outcomes (ABC-I and CGI-I). Table 1 presents the treatment ranking. For both outcomes, we reported the random-effects model. The findings of the fixed-effect model are presented in Supplementary Material 3. For the ABC-I network, we included 14 RCTs examining seven different interventions, including 1222 participants. We included 11 RCTs in the network for CGI-I, examining seven different interventions and involving 871 participants.

**Figure 2:**
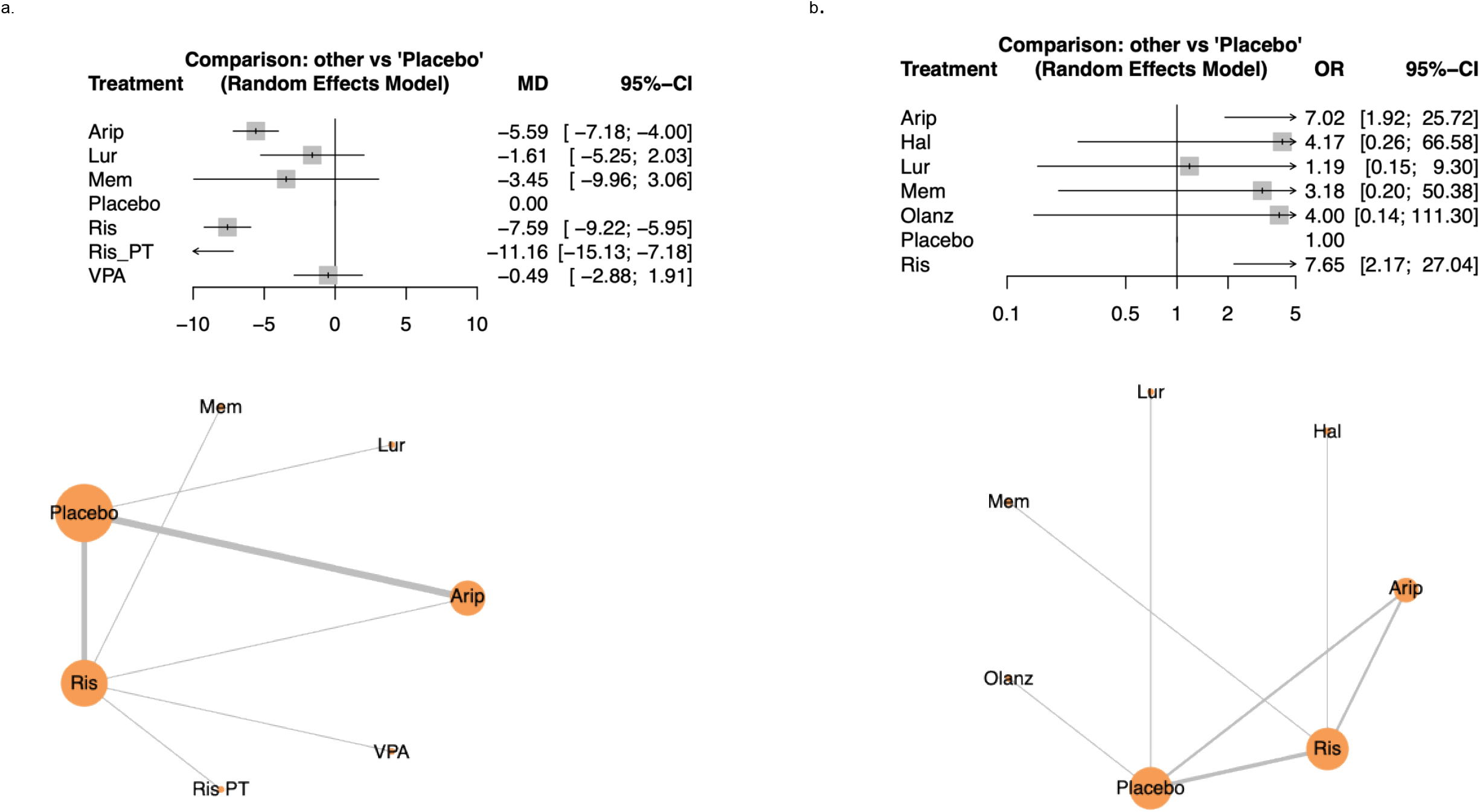
Clinical outcomes with a. representing ABC-I scores and b. representing CGI-I scores.

**Table 1:** Treatment ranking for primary and secondary outcomes.

| Interventions | P-scores |
| --- | --- |
| **Rank of interventions (Aberrant Behavior Checklist-Irritability score)** |  |
| Risperidone + parent training | 0.9917 |
| Risperidone | 0.8175 |
| Aripiprazole | 0.6231 |
| Memantine | 0.4546 |
| Lurasidone | 0.3069 |
| Valproic acid | 0.1916 |
| Placebo | 0.1146 |
| **Rank of interventions (Clinical Global Improvement-Improvement score)** |  |
| Risperidone | 0.7601 |
| Aripiprazole | 0.7236 |
| Haloperidol | 0.5588 |
| Olanzapine | 0.5491 |
| Memantine | 0.4904 |
| Lurasidone | 0.2502 |
| Placebo | 0.1678 |
| **Rank of interventions (Based on the highest number of dropouts due to adverse effects)** |  |
| Aripiprazole | 0.7186 |
| Risperidone + parent training | 0.6519 |
| Methylphenidate | 0.6387 |
| Placebo | 0.5263 |
| Risperidone | 0.2403 |
| Lurasidone | 0.2242 |
| **Rank of interventions (Based on the highest number of overall adverse effects recorded)** |  |
| Methylphenidate | 0.8371 |
| Risperidone | 0.6928 |
| Aripiprazole | 0.5187 |
| Lurasidone | 0.3942 |
| Placebo | 0.0572 |
| **Rank of interventions (Based on the highest number of sedations recorded)** |  |
| Valproic acid | 0.7851 |
| Risperidone | 0.6653 |
| Olanzapine | 0.6239 |
| Aripiprazole | 0.6197 |
| Risperidone + parent training | 0.5732 |
| Memantine | 0.3146 |
| Lurasidone | 0.3142 |
| Placebo | 0.1040 |
| **Rank of interventions (Based on the highest number of weight gain recorded)** |  |
| Olanzapine | 0.8288 |
| Valproic acid | 0.7582 |
| Risperidone | 0.7003 |
| Risperidone + parent training | 0.4698 |
| Lurasidone | 0.3946 |
| Aripiprazole | 0.2859 |
| Placebo | 0.0624 |

Compared with the placebo, the combined treatment with risperidone and parent training (PT) (MD=-11.16, 95% confidence interval [CI] =-15.13,-7.18), followed by risperidone (MD =-7.59, 95% CI =-9.22, -5.95) and aripiprazole (MD=-5.59, 95% CI= −7.18; −4) showed the largest effect on the ABC-I scores. These interventions were also at the top of the treatment ranking (Table 1). The 95% CIs for the ORs of lurasidone, divalproex sodium, and memantine crossed the no-effect line. Only 4.4% of the observed variability was attributed to heterogeneity (tau^2^ =0.17). Two RCTs in the current NMA involving aripiprazole, which included most participants, were conducted by the pharmaceutical company that manufactures aripiprazole.

Risperidone (OR = 7.65, 95% CI = 2.17; 27.04), followed by aripiprazole (OR = 7.02, 95% CI = 1.92; 25.72), produced the largest effect on CGI-I scores compared with placebo. The 95% CI of ORs for lurasidone, olanzapine, haloperidol and memantine included ‘1’. Risperidone and aripiprazole were also at the top of the treatment ranking (Table 1). The most observed variability (I2 = 75.4%) was attributed to heterogeneity (tau^2^ = 0.96). Sensitivity analysis by removing studies with high RoB (Miral et al., 2008; Nikvarz et al., 2016) did not substantially reduce heterogeneity. We systematically removed one study at a time and reanalysed data as an additional sensitivity analysis. We found that removing Marcus’s study reduced heterogeneity to I^2^ = 43.1% and tau^2^ = 0.32. Marcus’s study initially involved four different intervention arms based on different doses of aripiprazole. These data were combined for our analysis into a 2-arm intervention.

The sensitivity analysis also showed that the treatment ranking was not robust; the rank changed when data from any of the following sources were removed: Kent et al. (2013), McCracken et al. (2002), McDougle et al. (1998), Marcus et al. (2009), or Ghanizadeh et al. (2014). However, the ranking remained the same when the remaining studies were removed. As part of sensitivity analyses, we used the ‘netimpact’ function (Rücker et al., 2020) to assess the impact of the individual study in the pooled estimates and identified that four studies had the highest impact (Hollander et al., 2006; Miral et al., 2008; Nikvarz et al., 2017; Loebel et al., 2016) within the CGI-I NMA. However, these four studies each assessed only a specific medication (see supplementary material 3). Removing these studies removed their respective nodes in the network but did not change the overall treatment ranking of other interventions.

### Secondary outcomes

We pooled data on the overall adverse effects (AE) rate, weight gain, sedation, and dropout due to AE. Figure 3 shows the network’s structure and NMA results. The fixed-effects model results are presented in supplementary material 3.

**Figure 3.**
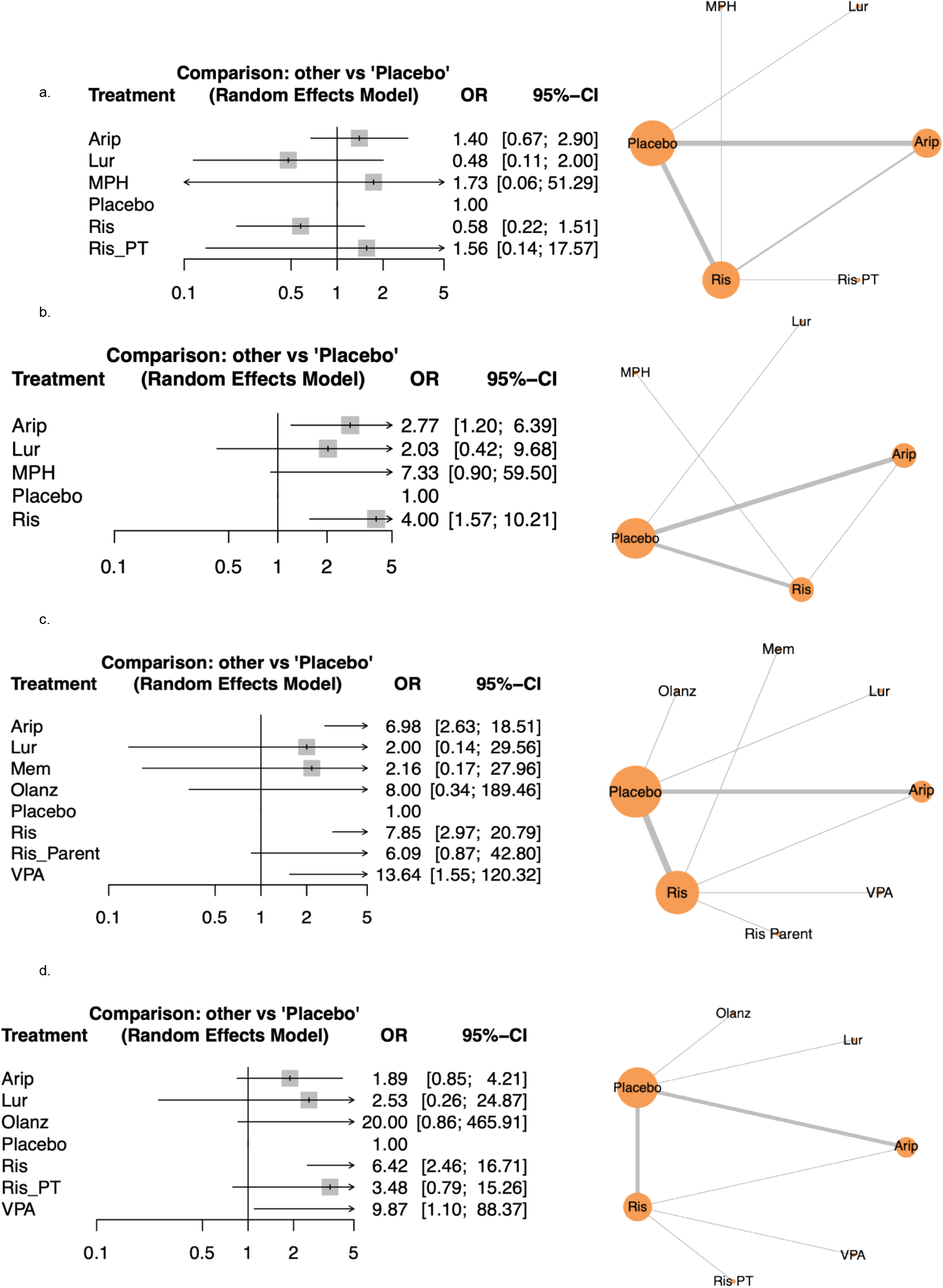
Safety/adverse events (AE) outcomes with a. representing drop out rates due to AE, b. representing overall AE, c. representing sedation and d. representing weight gain.

We included 15 RCTs on six interventions in the network, involving 1275 participants, to calculate the dropout rate due to AE. Lurasidone, methylphenidate, and the combined treatment of risperidone and PT had only one direct comparison. The finding of a dropout rate due to AE is unreliable because the 95% CIs for the ORs crossed the line of no effect for all interventions. There was no evidence of heterogeneity (tau^2^ = 0).

We included 12 studies in the network, with five different interventions and 993 participants, to estimate the overall rate of AEs. Methylphenidate showed the largest number of AEs compared with placebo (OR= 7.33, 95% CI= 0.90, 59.50), followed by risperidone (OR= 4.00, 95% CI= 1.57, 10.21), aripiprazole (OR= 2.77, 95% CI= 1.20, 6.39) and then, lurasidone (OR= 2.03, 95% CI= 0.42, 9.68), with 51.7% of the total observed variability attributed to heterogeneity (tau^2^ = 0.5). However, the ORs for lurasidone and methylphenidate’s credibility intervals included ‘1’.

We included 16 studies with eight different interventions in the network, involving 1257 participants, to calculate the sedation rate, one of the common AEs related to antipsychotic treatment. Divalproex sodium showed the highest OR (OR = 13.64, 95% CI = 1.55, 120.32), followed by olanzapine (OR = 8, 95% CI = 0.36, 189.46), risperidone (OR = 7.85, 95% CI = 2.97, 20.79), and aripiprazole (OR = 6.98, 95% CI = 2.63, 18.51). However, the 95% CI of lurasidone, memantine, combined risperidone and parent training, and olanzapine’s OR included ‘1’. Heterogeneity was low to moderate (I^2^= 44.2%, tau^2^ = 0.61).

We included 13 studies with seven different interventions in the network, involving 1161 participants, to calculate the rate of weight gain, a common antipsychotic-related AE. The odds of weight gain were greatest with divalproex sodium treatment (OR = 9.87, 95% CI = 1.10, 88.37), followed by risperidone (OR = 6.42, 95% CI = 2.46, 16.71). The ORs for combined risperidone and parent training, olanzapine, lurasidone, and aripiprazole, with 95% CIs, were wide and crossed the line of no effect. Heterogeneity was low (I^2^= 13%, tau^2^ = 0.13).

## Discussion

Network meta-analysis allows the pooling of evidence on multiple interventions from a set of RCTs, each comparing two or more interventions of interest. This provides a more inclusive approach than pairwise meta-analysis since all pairwise comparisons of treatments can be examined. If the effect estimates reveal substantial differences between treatments, a ranking of treatments can be useful in informing decisions for clinicians and policymakers. In this NMA, we explored both the efficacy and intervention-related adverse effects of antipsychotic medicines on autistic people of all ages. However, only one small study included adults, and the rest included children and adolescents.

In the current NMA, we analysed data from 22 RCTs involving 1562 autistic people. The data were available on five antipsychotics, namely risperidone, aripiprazole, haloperidol, olanzapine, and lurasidone, and three non-antipsychotic medicines, namely memantine, divalproex sodium, and methylphenidate. Two studies combined non-medicinal interventions, such as parent training and behavioural treatment using virtual reality, with risperidone and compared them with risperidone treatment alone. However, the data from only the RCT involving the combination of risperidone and parent training could be used for the current NMA.

As per the ORs, the combined intervention of the parent training and risperidone, risperidone alone and aripiprazole significantly improved ABC-I scores. This was reflected in the ranking. Conversely, there was no evidence of an effect of lurasidone, valproate, or memantine. Although ABC-I is supposed to assess irritability symptoms, this subsection of ABC includes 15 very different behaviours that may not be directly related to irritability. So, it cannot be concluded that improving the ABC-I score equates to an improvement in aggression or irritability. A recent NMA involving eight RCTs of antipsychotics showed that risperidone and aripiprazole significantly improved ABC-I scores in autistic children (Fallah et al., 2019). Although expected, we have, for the first time, shown that combining pharmacological therapy with a non-pharmacological intervention is better than pharmacological therapy alone. A recent systematic review found mild-to-moderate treatment effects of certain parent-training packages on core autism symptoms and associated behaviours (Deb et al., 2020).

According to the OR values, risperidone and aripiprazole showed the greatest improvement in the CGI-I score. There was no statistically significant difference in CGI-I scores between placebo and lurasidone, olanzapine, haloperidol and memantine. The ranking should not be overemphasised regarding CGI-I scores, given how close some relative effects are and the substantial impact on the sensitivity analyses performed for this outcome.

Clinicians need to weigh the efficacy against the risk of adverse effects in each case when choosing an intervention. National (NICE, 2015) and international guidelines (Deb et al., 2009) recommend using non-pharmacological interventions first for BtC in autistic people before considering any pharmacological intervention. None of the interventions has shown a significant difference in dropout rate due to the intervention compared with placebo. This finding is unreliable because of their ORs’ wide 95% CI. We have not presented ranking data for secondary outcomes, as they could be misleading due to overlapping effect estimates.

Methylphenidate, risperidone and aripiprazole showed significantly higher rates of any drug-related adverse effects. However, the difference between methylphenidate and placebo was not statistically significant. Previous RCTs of methylphenidate on autistic people (Joshi et al., 2021) and ID (Tarrant et al., 2018) did not show a statistically significant difference in the adverse effects rate between the methylphenidate and the placebo groups. However, in the current NMA, only one small RCT involving methylphenidate was included. A recent small NMA of antipsychotic RCTs in autistic children ranked risperidone and aripiprazole at the top for showing adverse effects (Fallah et al., 2019).

As for the rate of sedation, valproate, olanzapine, risperidone, and aripiprazole showed the highest ORs. The difference between olanzapine and placebo was not statistically significant. The same was true for lurasidone, memantine, and the combined treatment of risperidone and parent training. However, these effects were imprecisely estimated, as reflected in the wide 95% CIs. A recent NMA of the general population found olanzapine to have the highest risk ratio (RR) for sedation, followed by risperidone and aripiprazole, but none of these was statistically significant compared with placebo (Schneider-Thoma et al., 2022). Interestingly, the combination of risperidone and parent training did not show a statistically significant increase in sedation compared with placebo. This may be explained by the fact that, when combined with non-pharmacological interventions for BtC, a lower dose of risperidone may be required, leading to less sedation.

According to the OR, olanzapine, valproate and risperidone showed the most weight gain. The difference between olanzapine and placebo was not statistically significant. However, the effect measure for valproate is imprecise due to its very wide 95% CI. Aripiprazole, on the other hand, showed the smallest association in weight gain in the same direction, but its difference from placebo is not statistically significant. In a recent NMA of the general population, risperidone was associated with greater weight gain than aripiprazole (Kong et al., 2024).

The evidence is primarily based on autistic children, and there is not enough evidence available to draw any definitive conclusion about antipsychotic efficacy and tolerability for autistic adults. No RCT has properly assessed the effect of antipsychotics on psychiatric disorders like bipolar disorder or psychosis. No RCT assessed the participant’s quality of life, as symptom improvement does not necessarily equate to improving quality of life. Most RCTs were add-on trials of antipsychotics. Therefore, the confounding effect of concomitant medication and non-pharmacological interventions is unknown. Studies may not have used an optimum dose of antipsychotics. For example, a smaller dose may not be effective. On the other hand, a higher dose may produce adverse effects. There was not enough data available on the metabolic adverse effects and anticholinergic burden of antipsychotics.

The follow-up periods in the included studies were short, so the antipsychotics’ long-term effects are not known. Five RCTs (data not presented here) continued risperidone treatment for 48-52 months in an open-label design (see review by Deb, 2024). Most of these open-label studies showed sustained efficacy over time with reasonable tolerability. However, as no placebo was used, it is difficult to measure the placebo effect on the long-term efficacy and side effects of antipsychotics. Most studies were from the USA and were supported by pharmaceutical companies, although authors often claimed that the funders had no influence on their study design or findings. However, two major RCTs on aripiprazole, which included the largest sample sizes, were conducted by the pharmaceutical company that manufactures aripiprazole. Therefore, more independent research is needed to establish aripiprazole’s efficacy.

### Limitations

Certain limitations of this study have to be considered while interpreting data. The total number of RCTs involved is relatively small. As no data on IQ or the level of ID were presented in most papers, the transitivity assumption based on descriptive analysis may have remained unclear for this particular item. Not all RCTs included in the systematic review could be included in the NMA. Further properly powered RCTs are required to establish whether one antipsychotic is better than the other in improving symptoms of psychiatric disorders, BtC, and quality of life without compromising the tolerability. Properly powered RCTs combining antipsychotics with non-pharmacological interventions should be compared with non-pharmacological interventions alone.

## Data Availability

The main text and the supplementary material share most of the information. However, if any further information is needed, it will be provided upon reasonable request.

## Acknowledgements

The Imperial Biomedical Research Centre Facility, funded by the National Institute of Health Research (NIHR), UK, supported the study.

