## Supplementary material 1 for "A network meta-analysis of randomised controlled trials of antipsychotic medications to assess their comparative efficacy and tolerability in autistic people"

1) PRISMA NMA Checklist of Items to Include When Reporting A Systematic Review Involving a Network Meta-analysis

| Section/Topic | Item # | Checklist Item | Reported on Page # |
|---|---|---|---|
| TITLE | | | |
| Title | 1 | Identify the report as a systematic review *incorporating a network meta-analysis (or related form of meta-analysis).* | ***1*** |
| ABSTRACT | | | |
| Structured summary | 2 | Provide a structured summary including, as applicable:<br>**Background:** main objectives<br>**Methods:** data sources; study eligibility criteria, participants, and interventions; study appraisal; and *synthesis methods, such as network meta-analysis.*<br>**Results:** number of studies and participants identified; summary estimates with corresponding confidence/credible intervals; *treatment rankings may also be discussed. Authors may choose to summarize pairwise comparisons against a chosen treatment included in their analyses for brevity.*<br>**Discussion/Conclusions:** limitations; conclusions and implications of findings.<br>**Other:** primary source of funding; systematic review registration number with registry name. | 1 |
| INTRODUCTION | | | |
| Rationale | 3 | Describe the rationale for the review in the context of what is already known*, including mention of why a network meta-analysis has been conducted.* | ***1*** |
| Objectives | 4 | Provide an explicit statement of questions being addressed, with reference to participants, interventions, comparisons, outcomes, and study design (PICOS). | 1 |
| METHODS | | | |
| Protocol and registration | 5 | Indicate whether a review protocol exists and if and where it can be accessed (e.g., Web address); and, if available, provide registration information, including registration number. | 2 |
| Eligibility criteria | 6 | Specify study characteristics (e.g., PICOS, length of follow-up) and report characteristics (e.g., years considered, language, publication status) used as criteria for eligibility, giving rationale. *Clearly describe eligible treatments included in the treatment network, and note whether any have been clustered or merged into the same node (with justification).* | ***2*** |
| Information sources | 7 | Describe all information sources (e.g., databases with dates of coverage, contact with study authors to identify additional studies) in the search and date last searched. | 2 |

| | | | |
|---|---|---|---|
| Search | 8 | Present full electronic search strategy for at least one database, including any limits used, such that it could be repeated. | In appendix |
| Study selection | 9 | State the process for selecting studies (i.e., screening, eligibility, included in systematic review, and, if applicable, included in the meta-analysis). | 2 |
| Data collection process | 10 | Describe method of data extraction from reports (e.g., piloted forms, independently, in duplicate) and any processes for obtaining and confirming data from investigators. | 2 |
| Data items | 11 | List and define all variables for which data were sought (e.g., PICOS, funding sources) and any assumptions and simplifications made. | 2 |
| **Geometry of the network** | **S1** | Describe methods used to explore the geometry of the treatment network under study and potential biases related to it. This should include how the evidence base has been graphically summarized for presentation, and what characteristics were compiled and used to describe the evidence base to readers. | ***2*** |
| Risk of bias within individual studies | 12 | Describe methods used for assessing risk of bias of individual studies (including specification of whether this was done at the study or outcome level), and how this information is to be used in any data synthesis. | 2 |
| Summary measures | 13 | State the principal summary measures (e.g., risk ratio, difference in means). *Also describe the use of additional summary measures assessed, such as treatment rankings and surface under the cumulative ranking curve (SUCRA) values, as well as modified approaches used to present summary findings from meta-analyses.* | 3 |
| Planned methods of analysis | 14 | Describe the methods of handling data and combining results of studies for each network meta-analysis. This should include, but not be limited to:<br>• *Handling of multi-arm trials;*<br>• *Selection of variance structure;*<br>• *Selection of prior distributions in Bayesian analyses; and*<br>• *Assessment of model fit.* | 2-3 |
| **Assessment of Inconsistency** | **S2** | Describe the statistical methods used to evaluate the agreement of direct and indirect evidence in the treatment network(s) studied. Describe efforts taken to address its presence when found. | 2-3 |
| Risk of bias across studies | 15 | Specify any assessment of risk of bias that may affect the cumulative evidence (e.g., publication bias, selective reporting within studies). | 2-3 |
| Additional analyses | 16 | Describe methods of additional analyses if done, indicating which were pre-specified. This may include, but not be limited to, the following:<br>• Sensitivity or subgroup analyses;<br>• Meta-regression analyses;<br>• *Alternative formulations of the treatment network; and* | ***3*** |

- *Use of alternative prior distributions for Bayesian analyses (if applicable).*

RESULTS†

| | | | |
|---|---|---|---|
| Study selection | 17 | Give numbers of studies screened, assessed for eligibility, and included in the review, with reasons for exclusions at each stage, ideally with a flow diagram. | 3 |
| **Presentation of network structure** | **S3** | Provide a network graph of the included studies to enable visualization of the geometry of the treatment network. | ***In figures*** |
| **Summary of network geometry** | **S4** | Provide a brief overview of characteristics of the treatment network. This may include commentary on the abundance of trials and randomized patients for the different interventions and pairwise comparisons in the network, gaps of evidence in the treatment network, and potential biases reflected by the network structure. | ***4-5*** |
| Study characteristics | 18 | For each study, present characteristics for which data were extracted (e.g., study size, PICOS, follow-up period) and provide the citations. | In appendix |
| Risk of bias within studies | 19 | Present data on risk of bias of each study and, if available, any outcome level assessment. | 3 and in appendix |
| Results of individual studies | 20 | For all outcomes considered (benefits or harms), present, for each study: 1) simple summary data for each intervention group, and 2) effect estimates and confidence intervals. *Modified approaches may be needed to deal with information from larger networks.* | In appendix |
| Synthesis of results | 21 | Present results of each meta-analysis done, including confidence/credible intervals. *In larger networks, authors may focus on comparisons versus a particular comparator (e.g. placebo or standard care), with full findings presented in an appendix. League tables and forest plots may be considered to summarize pairwise comparisons.* If additional summary measures were explored (such as treatment rankings), these should also be presented. | In appendix |
| **Exploration for inconsistency** | **S5** | Describe results from investigations of inconsistency. This may include such information as measures of model fit to compare consistency and inconsistency models, *P* values from statistical tests, or summary of inconsistency estimates from different parts of the treatment network. | ***3-4 and in appendix*** |
| Risk of bias across studies | 22 | Present results of any assessment of risk of bias across studies for the evidence base being studied. | 3 and in appendix |
| Results of additional analyses | 23 | Give results of additional analyses, if done (e.g., sensitivity or subgroup analyses, meta-regression analyses*, alternative network geometries studied, alternative choice of prior distributions for Bayesian analyses,* and so forth). | ***3*** |

| | | | |
|---|---|---|---|
| **DISCUSSION** | | | |
| Summary of evidence | 24 | Summarize the main findings, including the strength of evidence for each main outcome; consider their relevance to key groups (e.g., healthcare providers, users, and policy-makers). | 6-7 |
| Limitations | 25 | Discuss limitations at study and outcome level (e.g., risk of bias), and at review level (e.g., incomplete retrieval of identified research, reporting bias). *Comment on the validity of the assumptions, such as transitivity and consistency. Comment on any concerns regarding network geometry (e.g., avoidance of certain comparisons).* | 7 |
| Conclusions | 26 | Provide a general interpretation of the results in the context of other evidence, and implications for future research. | 6 |
| **FUNDING** | | | |
| Funding | 27 | Describe sources of funding for the systematic review and other support (e.g., supply of data); role of funders for the systematic review. This should also include information regarding whether funding has been received from manufacturers of treatments in the network and/or whether some of the authors are content experts with professional conflicts of interest that could affect use of treatments in the network. | *8* |

PICOS = population, intervention, comparators, outcomes, study design.
* Text in italics indicateS wording specific to reporting of network meta-analyses that has been added to guidance from the PRISMA statement.
† Authors may wish to plan for use of appendices to present all relevant information in full detail for items in this section.

### 2) Search Terms

Supplementary Appendix: Search terms.

*Antipsychotics*
*atypical antipsychotics OR second generation antipsychotics OR new generation antipsychotics OR antipsychotic OR aripiprazole OR quetiapine OR olanzapine OR risperidone OR clozapine OR old generation antipsychotics OR typical antipsychotics OR first generation antipsychotics OR second generation antipsychotics OR chlorpromazine OR haloperidol OR paliperidone OR asenapine OR ziprasidone OR lurasidone OR cariprazine.*

AND

*ASD descriptors:*

child developmental disorder* OR pervasive developmental disorder* OR autis* OR PDD* OR ASD* OR Kanner* OR Asperger* OR Asperger* syndrome OR autism spectrum disorder OR Rett Syndrome OR childhood schizophrenia OR Fragile X syndrome OR neurodevelopmental disorder* OR NDD*.

AND

*Outcome:*

Psychosis OR schizophrenia OR hallucination OR delusion OR mania OR hypomania OR autism core symptoms OR ASD core symptoms OR ASD symptoms OR autism symptoms OR social interaction OR communication problems OR social communication OR agitation OR irritability OR aggression OR behavioural problems OR problem behaviors OR challenging behaviour OR behaviour* that challenge OR behaviour of concern OR maladaptive behaviour OR disruptive behaviour OR disturbed behaviour OR distressed behaviour OR stereotypy OR restricted behaviour OR repetitive patterns of behaviour OR restricted interests OR restrictive activities OR social communication OR repetitive behaviour OR communication* OR inattention OR hyperactivity OR insistence on sameness OR sameness OR sleep problem OR insomnia OR self injurious behaviour OR self-mutilation OR temper tantrum OR tantrum OR aggression to others OR aggression to property OR sexual aggression OR sexual deviance OR mental state OR global improvement OR quality of life OR CGI.

AND

*RCT:*

clinical trial* OR randomization* OR randomisation OR research design OR randomized controlled trial OR randomi#ed control* trial* OR RCT OR controlled clinical trial OR double-blind procedure OR random* OR trial* OR control* OR blind* OR crossover OR crossover procedure OR crossover trial* OR volunteer* OR placebo* OR randomly OR control* OR ((singl* or doubl* or trebl* or tripl*) adj3 (blind* or mask*)) OR comparative stud* OR psychopharmacology AND not (animal OR nonhuman) treatment OR effectiveness evaluation OR treatment outcomes OR follow-up studies OR evaluat* adj3 stud*.

3) Search strategy

ID Search Hits
#1 ("atypical antipsychotics"):ti,ab,kw (Word variations have been searched) 1879
#2 ("second generation antipsychotics"):ti,ab,kw (Word variations have been searched) 639
#3 ("new generation antipsychotics"):ti,ab,kw (Word variations have been searched) 25
#4 ("antipsychotic"):ti,ab,kw (Word variations have been searched) 11915
#5 ("aripiprazole"):ti,ab,kw (Word variations have been searched) 1851
#6 ("quetiapine"):ti,ab,kw (Word variations have been searched) 2117
#7 ("olanzapine"):ti,ab,kw (Word variations have been searched) 3955
#8 ("risperidone"):ti,ab,kw (Word variations have been searched) 3872
#9 ("clozapine"):ti,ab,kw (Word variations have been searched) 1638
#10 ("old generation antipsychotics"):ti,ab,kw (Word variations have been searched) 0
#11 ("typical antipsychotics"):ti,ab,kw (Word variations have been searched) 225
#12 ("first generation antipsychotics"):ti,ab,kw (Word variations have been searched) 154
#13 (second generation antipsychotics):ti,ab,kw (Word variations have been searched)836
#14 ("chlorpromazine"):ti,ab,kw (Word variations have been searched) 1321
#15 ("haloperidol"):ti,ab,kw (Word variations have been searched) 3391
#16 ("paliperidone"):ti,ab,kw (Word variations have been searched) 724
#17 ("asenapine"):ti,ab,kw (Word variations have been searched) 202
#18 ("ziprasidone"):ti,ab,kw (Word variations have been searched) 792
#19 ("lurasidone"):ti,ab,kw (Word variations have been searched) 548
#20 ("cariprazine"):ti,ab,kw (Word variations have been searched) 230
#21 #1 OR #2 OR #3 OR #4 OR #5 OR #6 OR #7 OR #8 OR #9 OR #10 OR #11 OR #12 OR #13 OR #14 OR #15 OR #16 OR #17 OR #18 OR #19 OR #20 19327
#22 MeSH descriptor: [Developmental Disabilities] explode all trees 846
#23 (child developmental disorder*):ti,ab,kw (Word variations have been searched) 2857
#24 (pervasive developmental disorder*):ti,ab,kw (Word variations have been searched) 415
#25 (autis*):ti,ab,kw (Word variations have been searched) 5743
#26 (PDD*):ti,ab,kw (Word variations have been searched) 697
#27 (ASD*):ti,ab,kw (Word variations have been searched) 3984
#28 (Kanner*):ti,ab,kw (Word variations have been searched) 8
#29 (Asperger*):ti,ab,kw (Word variations have been searched) 303
#30 (asperger* syndrome):ti,ab,kw (Word variations have been searched) 243
#31 MeSH descriptor: [Asperger Syndrome] explode all trees 87
#32 MeSH descriptor: [Autism Spectrum Disorder] explode all trees 2722
#33 MeSH descriptor: [Rett Syndrome] explode all trees 67
#34 ("childhood schizophrenia"):ti,ab,kw (Word variations have been searched) 9
#35 ("Fragile X syndrome"):ti,ab,kw (Word variations have been searched) 199
#36 (neurodevelopmental disorder*):ti,ab,kw (Word variations have been searched) 1451
#37 (NDD*):ti,ab,kw (Word variations have been searched) 361
#38 #22 OR #23 OR #24 OR #25 OR #26 OR #27 OR #28 OR #29 OR #30 OR #31 OR #32 OR #33 OR #34 OR #35 OR #36 OR #37 11597
#39 (Psychosis OR schizophrenia OR hallucination OR delusion):ti,ab,kw (Word variations have been searched) 26052
#40 (mania OR hypomania):ti,ab,kw (Word variations have been searched) 3023
#41 ("autism core symptoms" OR "ASD core symptoms" OR "ASD symptoms" OR "autism symptoms"):ti,ab,kw (Word variations have been searched) 297
#42 ("social interaction" OR "communication problems" OR "social communication"):ti,ab,kw (Word variations have been searched) 5866

#43 (agitation OR irritability OR aggression OR "behavioural problems" OR "problem behaviors"):ti,ab,kw (Word variations have been searched) 40073
#44 ("challenging behaviour" OR "behaviour of concern" OR "maladaptive behaviour" OR "disruptive behaviour" OR "disturbed behaviour" OR "distressed behaviour"):ti,ab,kw (Word variations have been searched) 1937
#45 (behaviour* NEAR challenge):ti,ab,kw (Word variations have been searched) 456
#46 ("stereotypy" OR "restricted behaviour" OR "repetitive patterns of behaviour" OR "restricted interests" OR "restrictive activities" OR "repetitive behaviour"):ti,ab,kw (Word variations have been searched) 946
#47 (repetitive NEAR behavi*r):ti,ab,kw (Word variations have been searched)635
#48 (communication*):ti,ab,kw (Word variations have been searched)28801
#49 (inattention OR hyperactivity OR sameness):ti,ab,kw (Word variations have been searched) 144660
#50 ("insistence on sameness"):ti,ab,kw (Word variations have been searched) 2
#51 ("sleep problem" OR "insomnia"):ti,ab,kw (Word variations have been searched) 16695
#52 ("self injurious behaviour" OR "self-mutilation" OR "temper tantrum" OR "tantrum" OR "aggression to others" OR "aggression to property" OR "sexual aggression" OR "sexual deviance" OR "mental state"):ti,ab,kw (Word variations have been searched) 10622
#53 ("global improvement" OR "quality of life" OR "CGI"):ti,ab,kw (Word variations have been searched) 175491
#54 #39 OR #40 OR #41 OR #42 OR #43 OR #44 OR #45 OR #46 OR #47 OR #48 OR #49 OR #50 OR #51 OR #52 OR #53 399811
#55 ((clinical trial*) OR (randomization*)):ti,ab,kw (Word variations have been searched) 905975
#56 ("randomisation" OR "research design" OR "randomized controlled trial"):ti,ab,kw (Word variations have been searched) 1369437
#57 (randomi*ed NEXT control* NEXT trial*):ti,ab,kw (Word variations have been searched) 718886
#58 ("RCT" OR "controlled clinical trial" OR "double-blind procedure"):ti,ab,kw (Word variations have been searched) 303793
#59 (random*):ti,ab,kw (Word variations have been searched) 1368054
#60 (trial*):ti,ab,kw (Word variations have been searched) 1179614
#61 (control*):ti,ab,kw (Word variations have been searched) 1358312
#62 (blind*):ti,ab,kw (Word variations have been searched) 481846
#63 ("crossover" OR "crossover procedure"):ti,ab,kw (Word variations have been searched) 119018
#64 (crossover NEXT trial*):ti,ab,kw (Word variations have been searched) 17947
#65 (volunteer*):ti,ab,kw (Word variations have been searched) 86647
#66 (placebo*):ti,ab,kw (Word variations have been searched) 396422
#67 (control*):ti,ab,kw (Word variations have been searched) 1358312
#68 (randomly):ti,ab,kw (Word variations have been searched) 1285976
#69 ((singl* OR doubl* OR trebl* OR tripl*) NEAR (blind* OR mask*)):ti,ab,kw (Word variations have been searched) 432812
#70 (comparative stud*):ti,ab,kw (Word variations have been searched) 786111
#71 ((psychopharmacology) NOT (animal OR nonhuman treatment)):ti,ab,kw (Word variations have been searched) 2213
#72 ("effectiveness evaluation" OR "treatment outcomes" OR "follow-up studies"):ti,ab,kw (Word variations have been searched) 283646
#73 (evaluat* NEAR stud*):ti,ab,kw (Word variations have been searched) 203553
#74 #55 OR #56 OR #57 OR #58 OR #59 OR #60 OR #61 OR #62 OR #63 OR #64 OR #65 OR #66 OR #67 OR #68 OR #69 OR #70 OR #71 OR #72 OR #73 1872252

#75 #21 AND #38 AND #54 AND #74 with Cochrane Library publication date Between May 2022 and Sep 2024 40

### 4) Eligibility criteria and data extraction form

#### Eligibility criteria

| | | | |
|---|---|---|---|
| Citation: | | | |
| Reviewer's initials: | | | |
| Date of scoring: | | | |
| Study design: Is the study a randomized controlled trial? | Y | N | U |
| Intervention: Does the intervention involve antipsychotics? | Y | N | U |
| Population: Do all participants have ASD (defined using a standardised method)? | Y | N | U |
| Is the control group matched? | Y | N | U |
| Is the control group unmatched? | Y | N | U |
| Outcome: Are the outcome measures repeatable? | Y | N | U |
| If all yes, include it for review. | Y | N | U |
| If uncertain get the full paper for further check. | Y | N | U |
| If not all yes and no uncertainty exclude. | Y | N | U |

Decision:

Y: yes; N: No; U: uncertain.

Reason for exclusion:

Data extraction proforma (adapted from Cochrane Collaboration template)

Notes on using data extraction form:

- Be consistent in the order and style you use to describe the information for each report.
- Record any missing information as unclear or not described, to make it clear that the information was not found in the study report(s), not that you forgot to extract it.
- Include any instructions and decision rules on the data collection form, or in an accompanying document. It is important to practice using the form and give training to any other authors using the form.

Title of the systematic review:

General Information

| |
|---|
| Date form completed *(dd/mm/yyyy)* |
| Name/ID of person extracting data |
| Reference citation (full citation) |
| Study author contact details (Email) |
| Publication type *(e.g., full report, abstract, letter)* |
| Notes: |

*Characteristics of the included study*

Participants

| | Description<br>*Include comparative information for each intervention or comparison group if available* |
|---|---|
| Population description *(from which study participants are drawn)* | |
| Setting *(e.g., intensive care unit, service providers, institutions, day care centre etc)* | |
| Method of recruitment of participants *(e.g., phone, mail, clinic patients)* | |
| Informed consent obtained | Yes No Unclear |
| Intervention group | Age of participants (range, mean & SD) |
| | Number (%) of participants by gender |
| | Number (%) with ID, ADHD or other NDDs |
| | Type of pharmacological regime (name of the antipsychotic) and the dose |
| | Co morbidity (psychiatric) |
| | Co morbidity (physical) |

| | |
|---|---|
| | Adverse events (number and %) |
| Control group | Age of participants (range, mean & SD) |
| | Number (%) of participants by gender |
| | Number (%) with ID, ADHD or other NDDs |
| | Type of pharmacological regime (placebo or another medication) + name + dose |
| | Co morbidity (psychiatric) |
| | Co morbidity (physical) |
| | Adverse events (number and %) |

Methods

| | Descriptions as stated in report/paper | Location in text or source *(page & ¶/fig/table/other)* |
|---|---|---|
| Aim of study *(e.g., efficacy, equivalence, pragmatic)* | | |
| Design *(e.g., parallel, crossover)* | | |
| Sampling technique (e.g., random) | | |
| Method of establishing ASD diagnosis (if known) (clinical or ICD or DSM or ADI-R or ADOS etc.) | | |

Outcomes

*Copy and paste table for each outcome.*

| | Description as stated in report/paper | | | | Location in text or source *(page & ¶/fig/table/other)* |
|---|---|---|---|---|---|
| Primary outcome if dichotomous (e.g., %) (name the outcome and the instrument used to measure the outcome) | Number (%) in the intervention arm | Total number of participants in the intervention arm | Number (%) in the control arm | Total number of participants in the control arm | |

| | | | | |
|---|---|---|---|---|
| Primary outcome if continuous | Mean in the intervention arm (95% CI) | SD in the intervention arm (95% CI) | Mean in the control arm (95% CI) | SD in the control arm (95% CI) |
| Duration of intervention (weeks/months) (if crossover, add duration of baseline and washout period) | | | | |
| Duration of follow up (weeks/months) | | | | |
| Statistical methods used and appropriateness of these *(e.g., proportion, %, risk ratio, odds ratio)* | | | | |
| Secondary outcomes | | | | |
| Number of missing data | | | | |
| Reason for missing data | | | | |
| Other | | | | |
| Is outcome/tool validated? | Yes No Unclear | | Name of the tool: | |
| Notes: | | | | |

Other information

| | Description as stated in report/paper | Location in text or source *(page & ¶/fig/table/other)* |
|---|---|---|
| Main findings (statistically significant difference or not; provide P value or other relevant data in support of main findings (primary and secondary outcomes) | | |

| |
|---|
| Key conclusions of study authors |
| Your critique of the study (any design flaw etc.) |
| Your own overall conclusion |
| Correspondence required for further study information *(from whom, what and when)* |
| Notes: |

Other

| |
|---|
| Study funding sources *(including role of funders)* |
| Possible conflicts of interest *(for study authors)* |
| Notes: |

5) Reasons for exclusion of studies

| Citation | Reasons for exclusion |
|---|---|
| Aman MG, Kasper W, Manos G, et al: Line-item analysis of the aberrant behavior checklist: Results from two studies of aripiprazole in the treatment of irritability associated with autistic disorder. J Child Adolesc Psychopharmacol 2010; 20(5):415-422. | This paper has included data from two aripiprazole papers (Owen 2009 + Marcus 2009), and we have already included these two papers separately in primary studies. |
| Anderson LT, Campbell M, Adams P, et al: The effects of haloperidol on discrimination learning and behavioral symptoms in autistic children. J Autism Dev Disord 1989; 19(2):227-239. | This study is on haloperidol, which is not a second-generation APT. We included new-generation antipsychotic RCTs only. |
| Anderson LT, Campbell M, Grega DM, et al: Haloperidol in the treatment of infantile autism: Effects on learning and behavioral symptoms. Am J Psychiatry 1984; 141(10):1195–1202. | This study is on haloperidol, which is not a second-generation APT. We included new-generation antipsychotic RCTs only. |
| Benton TD. Aripiprazole to treat irritability associated with autism: A placebo-controlled, fixed-dose trial. Curr Psychiatry Rep 2011; 13(2):77-79. | Same data as in Marcus's 2009 aripiprazole study, which is already included. |
| Caicedo C, Williams SH. Risperidone improves behavior in children with autism. J Fam Pract 2002; 51(11):915. | It is a summary abstract of the McCracken (RUPP) 2002 RCT on risperidone, which is already included. |
| Campbell M, Anderson LT, Small AM, et al: The effects of haloperidol on learning and behavior in autistic children. J Autism Dev Disord 1982; 12(2):167-175. | This study is on haloperidol, which is not a second-generation APT. We included new-generation antipsychotic RCTs only. |
| Cohen IL, Campbell M, Posner D. A study of haloperidol in young autistic children: a within-subjects design using objective rating scales. Psychopharmacol Bull 1980; 16(3):63-65. | This study is on haloperidol, which is not a second-generation APT. We included new-generation antipsychotic RCTs only. |
| Cohen IL, Campbell M, Posner D. Behavioral effects of haloperidol in young autistic children. An objective analysis using a within-subjects reversal design. J Am Acad Child Psychiatry 1980; 19(4):665-677. | This study is on haloperidol, which is not a second-generation APT. We included new-generation antipsychotic RCTs only. |
| EUCTR2006-005346-37-NL. A randomized, double-Blind, placebo-controlled maintenance of effect study of olanzapine in the treatment of disruptive behavioral symptoms in children and adolescents with Pervasive Developmental Disorders 2006. https://www.cochranelibrary.com/central/doi/10.1002/central/CN-01892127/full (last accessed on 01.12.22) | Results are not available on the Clinical Trial website. |

| | |
|---|---|
| EUCTR2015-001320-31-Outside-EU/EEA. A study to evaluate the efficacy and safety of risperidone (R064766) in children and adolescents with irritability associated with autistic disorder. https://www.cochranelibrary.com/central/doi/10.1002/central/CN-01870569/full 2015 (last accessed on 01.12.22) | Results are not available on the Clinical Trial website. |
| Findling RL, Mankoski R, Timko K, et al: A randomized controlled trial investigating the safety and efficacy of aripiprazole in the long-term maintenance treatment of pediatric patients with irritability associated with autistic disorder. J Clin Psychiatry 2014; 75(1):22-30. | It is a placebo-controlled withdrawal study. We excluded withdrawal studies. |
| Hellings JA, Zarcone JR, Crandall K, et al: Weight gain in a controlled study of risperidone in children, adolescents and adults with mental retardation and autism. J Child Adolesc Psychopharmacol 2001; 11(3):229-238. | This paper presented secondary data, but there is no reference to the primary study. |
| Hellings JA, Zarcone JR, Reese RM, et al: A crossover study of risperidone in children, adolescents and adults with mental retardation. J Autism Dev Disord 2006; 36(3):401-411. | This is a cross-over study, and no specific data for Phase I, are available, which is an inclusion criterion in our protocol. |
| Johnson & Johnson Pharmaceutical Research & Development, L.L.C. Risperidone in the treatment of children and adolescents with autistic disorder: A double-blind, placebo-controlled study of efficacy and safety, followed by an open-label extension study of safety. clinicaltrials.gov; 2014. https://clinicaltrials.gov/ct2/show/NCT00576732 (last accessed on 01.12.22) | The same data are presented in the Kent 2013 paper, which is already included. |
| Lamberti M, Siracusano R, Italiano D, et al : Head-to-head comparison of aripiprazole and risperidone in the treatment of ADHD symptoms in children with autistic spectrum disorder and ADHD: a pilot, open-label, randomized controlled study. Paediatr Drugs 2016; 18(4):319-329. | The outcomes are more related to ADHD than ASD. Also, the study is not blinded. |
| Levine SZ, Kodesh A, Goldberg Y, et al: Initial severity and efficacy of risperidone in autism: Results from the RUPP trial. Eur Psychiatry 2016; 32:16-20. | The same data was presented in the RUPP/McCracken 2002 study. |
| Martsenkovsky I, Martsenkovska I, Martsenkovskyi D. Risperidon and atomoxetine in the treatment of several and challenging behaviors in children with PDD. Eur Psychiatry 2015; 30:195. | It is a conference abstract, and there are no data on the number of children included in the placebo, risperidone and atomoxetine groups. Also, the treatment is for ADHD rather than ASD. |

| | |
|---|---|
| McCracken. Risperidone treatment of autistic disorder: Longer-term benefits and blinded discontinuation after 6 months. Am J Psychiatry 2005; 162(7):1361-1369. | It is a placebo-controlled discontinuation study. |
| McCracken JT, McGough J, Shah B, et al: Risperidone was safe and effective for short term treatment of children with autism and serious behavioural disturbances. Evid Based Med 2003; 8(1):22. | It is a summary report of the McCracken/RUPP 2002 study. |
| NCT00005014. Treatment of autism in children and adolescents. 2000. https://www.cochranelibrary.com/central/doi/10.1002/central/CN-02024836/full (last accessed on 01.12.22) | Same data as in other secondary RUPP 2002 papers; Aman et al., 2005, Anderson et al., 2007, Aman et al., 2008, Arnold et al., 2010, Levine et al., 2016, Lindsay et al., 2006, McCracken et al., 2002, which are already included in our review. |
| NCT00057408. A controlled study of olanzapine in children with autism. 2003. https://www.cochranelibrary.com/central/doi/10.1002/central/CN-01509239/full (last accessed on 01.12.22) | Results are not available on the Clinical Trial website. |
| NCT00870727. Study of aripiprazole in the treatment of pervasive developmental disorders. 2009. https://www.cochranelibrary.com/central/doi/10.1002/central/CN-01596587/full (last accessed on 01.12.22) | Results are not available on the Clinical Trial website. |
| NCT01171937. 2010 Risperidone treatment in children with autism spectrum disorder and high levels of repetitive behavior. https://www.cochranelibrary.com/central/doi/10.1002/central/CN-01530768/full (last accessed on 01.12.22). | Results are not available on the Clinical Trial website. |
| NCT01333072. Biomarkers in Autism of Aripiprazole and Risperidone Treatment (BAART). https://clinicaltrials.gov/show/NCT01333072 2010; (last accessed on 01.12.22). | It is the same as the De Vane et al., 2019 paper, which is already included in our review. |
| NTR294. Risperidone in children and adolescents with severe disruptive behavior problems. https://www.cochranelibrary.com/central/doi/10.1002/central/CN-01826301/full, 2005 (last accessed on 01.12.22) | No results were posted on the website. |
| CN138178/NCT00332241. A multicenter double-blind, randomized, placebo-controlled, flexible-dosed, parallel-group study of aripiprazole in the treatment of children and adolescents with autistic disorder. 2022. https://clinical-trials.otsuka.com/postings/cn138178 (last accessed on 01.12.22) | It is the same as the Owen et al., 2009 aripiprazole RCT that we have already included in our review. |

| | |
|---|---|
| Remington G, Sloman L, Konstantareas M, et al: Clomipramine versus haloperidol in the treatment of autistic disorder: A double-blind, placebo-controlled, crossover study. J Clin Psychopharmacol 2001; 21(4):440-444. | This study is on haloperidol which is not a new-generation APT. We have included only the new-generation antipsychotic RCTs in our review. |
| Stigler K, Wang Y, McDonald B, et al: Effects of aripiprazole on brain circuitry in youth with pervasive developmental disorders. Neuropsychopharmacol 2010; 35:S367. | This is only a conference abstract on neuroimaging outcomes. |
| Sunovion. A 6-week, randomized, parallel, double-blind, placebo-controlled, fixed-dose, multicenter study to evaluate the efficacy and safety of lurasidone in children and adolescent subjects with irritability associated with autistic disorder. 2016. https://clinicaltrials.gov/ct2/show/NCT01911442 (last accessed on 01.12.22) | This paper presented the same data as in the Loebel et al., 2016 paper, which is already included. |
| Troost PW, Lahuis BE, Steenhuis MP, et al: Long-term effects of risperidone in children with autism spectrum disorders: a placebo discontinuation study. J Am Acad Child Adolesc Psychiatry 2005; 44(11):1137-1144. | It is a discontinuation study. |
| Troost PW, Althaus M, Lahuis BE, et al: Neuropsychological effects of risperidone in children with pervasive developmental disorders: A blinded discontinuation study. J Child Adolesc Psychopharmacol 2006; 16(5):561-573. | It is a discontinuation study. |

Updated search

| | |
|---|---|
| Alsayouf H, Talo H. Risperidone and Aripiprazole in Children with Autism Spectrum Disorder Substantially Improves Core Signs and Symptoms in Combination with Standard Supportive Therapies: A Large, Single-Center, Retrospective Case Series. | This study was not an RCT, it was a review of case series. |
| Berloffa S, Masi G, Falcone F, Simonelli V, Narzisi A, Valente E, Viglione V, Milone A, Sesso G. Clozapine treatment for aggressive behaviors in youths with neurodevelopmental disorders. Journal of child and adolescent psychopharmacology. 2024 Apr 1;34(3):148-56. | This study is not an RCT but an observational study. |
| CTRI/2024/06/069237. Comparison of Propranolol versus Risperidone in children with Autism Spectrum Disorder. 2024. https://www.cochranelibrary.com/central/doi/10.1002/central/CN-02722183/full (last accessed on 20.09.24) | This was a protocol abstract with no results. |

| | |
|---|---|
| Ebrahimi P, Seyedmirzaei H, Moradi K, Bagheri S, Moeini M, Mohammadi MR, Akhondzadeh S. Cilostazol as adjunctive therapy in treatment of children with autism spectrum disorders: a double-blind and placebo-controlled randomized trial. International clinical psychopharmacology. 2023 Mar 1;38(2):89-95. | It is a study assessing cilostazol vs combination (COMB) treatment of Risperidone and placebo. Studies with COMB interventions was excluded. |
| Hellings J. Pharmacotherapy in autism spectrum disorders, including promising older drugs warranting trials. World Journal of Psychiatry. 2023 Jun 6;13(6):262. | This was a review paper. |
| Hermans RA, Ringeling LT, Liang K, Kloosterboer SM, de Winter BC, Hillegers MH, Koch BC, Dierckx B. The effect of therapeutic drug monitoring of risperidone and aripiprazole on weight gain in children and adolescents: the SPACe 2: STAR (trial) protocol of an international multicentre randomised controlled trial. BMC psychiatry. 2022 Dec 20;22(1):814. | It is a protocol paper with no results available. |
| Hodgins GE, Winsor K, Barnhill J. Pharmacotherapy of Disruptive Behaviors in Children with Intellectual Disabilities. Pediatric Drugs. 2022 Sep;24(5):465-82. | This was a review paper. |
| Iffland M, Livingstone N, Jorgensen M, Hazell P, Gillies D. Pharmacological intervention for irritability, aggression, and self-injury in autism spectrum disorder (ASD). Cochrane database of systematic reviews. 2023(10). | This was a review paper. |
| IRCT20090117001556N156. Metformin as an adjuvant therapy for autism. 2024. https://www.cochranelibrary.com/central/doi/10.1002/central/CN-02687270/full (last accessed on 20.09.24). | This was another COMB treatment and a protocol paper with no results. |
| Kim BU, Kim HW, Park EJ, Kim JH, Boon-Yasidhi V, Tarugsa J, Reyes A, Manalo SG, Joung YS. Long-Term Improvement and Safety of Aripiprazole for Irritability and Adaptive Function in Asian Children and Adolescents with Autistic Disorder: A 52-Week, Multinational, Multicenter, Open-Label Study. Journal of child and adolescent psychopharmacology. 2022 Sep 1;32(7):390-9. | It is not an RCT but a prospective observational case study. |
| Mao AR. Psychopharmacology Management: Mood Instability and Impulsive Aggression in Youth With ASD. J Am Acad Child Adolesc Psychiatry 2022; 61: S21. | It is a review. |

| | |
|---|---|
| Martinez VM, Beato-Fernández L, Segura-Escobar E. Antipsychotics for irritability in children with Autism Spectrum Disorders. European Psychiatry. 2022 Jun;65(S1):S419-20. | It was not an RCT study. |
| Martsenkovsky I, Makarenko G, Martsenkovska I. Efficacy and safety of levetiracetam and risperidone for aggression irritability, and hyperactivity in adolescent with autism spectrum disorder. European Psychiatry. 2020 Jul 2;63. | This was a conference abstract with limited information. |
| Nasiri M, Parmoon Z, Farahmand Y, Moradi A, Farahmand K, Moradi K, Basti FA, Mohammadi MR, Akhondzadeh S. l-carnitine adjunct to risperidone for treatment of autism spectrum disorder-associated behaviors: A randomized, double-blind clinical trial. International Clinical Psychopharmacology. 2024 Jul 1;39(4):232-9. | This study was another COMB study, which we excluded. |
| NCT05491720. Comparable Efficacy of Transcranial Direct Current Stimulation and Pharmacological Treatments in Children With Autism Spectrum Disorder. 2022. https://clinicaltrials.gov/study/NCT05491720 (last accessed on 20.09.24) | No results available. |
| NCT05868720. Effect of risperidone vs aripiprazole on oxidative stress in patients with autism spectrum disorder: a randomized controlled trial. 2023. https://www.cochranelibrary.com/central/doi/10.1002/central/CN-02563046/full (last accessed on 20.09.24) | No results available. |
| NCT06315465. Efficacy of MAD as add-on Therapy in Comparison With Standard of Care in Children With ASD. 2024. https://www.cochranelibrary.com/central/doi/10.1002/central/CN-02681723/full (last accessed on 20.09.24) | Does not meet eligibility criteria (No antipsychotic med). |
| NL-OMON23089. Safety and pharmacokinetics of antipsychotics in children 2: studying TDM in an RCT. 2021. https://www.cochranelibrary.com/central/doi/10.1002/central/CN-02438766/full (last accessed on 20.09.24) | No control group or results available. |
| Rossignol DA, Frye RE. Psychotropic Medications for Sleep Disorders in Autism Spectrum Disorders. InHandbook of Autism and Pervasive Developmental Disorder: Assessment, Diagnosis, and Treatment 2022 | This was a review paper. |

| | |
|---|---|
| Aug 12 (pp. 1191-1217). Cham: Springer International Publishing. | |
| Salpekar JA, Scahill L. Psychopharmacology Management in Autism Spectrum Disorder. Pediatric Clinics. 2024 Apr 1;71(2):283-99. | This was a review paper. |
| Shakibaei F, Jelvani D. Effect of adding l-carnitine to risperidone on behavioral, cognitive, social, and physical symptoms in children and adolescents with autism: a randomized double-blinded placebo-controlled clinical trial. Clinical Neuropharmacology. 2023 Mar 1;46(2):55-9. | This study was another COMB study, which we excluded. |
| Sun C, Temelie A, Goulding H, Clark C, Yabs M, Fabian T. Long-Acting Injectable Antipsychotic Initiation in Child and Adolescent Patients with Psychiatric Disorders. Journal of Child and Adolescent Psychopharmacology. 2024 Aug 26. | It is a retrospective chart review not an RCT. |
| TMM D'A. Low-dose Abilify RTM and the treatment of disruptive behaviors in children with autism spectrum disorders. https://www.cochranelibrary.com/central/doi/10.1002/central/CN-02416350/full (last accessed on 20.09.24). | No paper/link to the actual dissertation/ thesis. |
| Veenstra-VanderWeele J. Assessment and Management of Irritability and Agitation in ASD. J Am Acad Child Adolesc Psychiatry 2022; 61: S139-S140. | It is a systematic review. Not RCT. |
| Williamson A, Raja V, Bailey L. Clozapine for Treatment Resistant Aggression in Autism. BJPsych Open. 2022 Jun;8(S1):S127-. | This is a case study and not RCT. |
| Yeung PP, Johnson KA, Riesenberg R, Orejudos A, Riccobene T, Kalluri HV, Malik PR, Varughese S, Findling RL. Cariprazine in pediatric patients with autism spectrum disorder: results of a pharmacokinetic, safety and tolerability study. Journal of Child and Adolescent Psychopharmacology. 2023 Aug 1;33(6):232-42. | It is an open label, multi-dose study without any placebo comparison. |
| Yeung P, Johnson KA, Riesenberg R, Orejudos A, Riccobene T, Kalluri HV, Malik P, Varughese S, Findling RL. Safety, Tolerability, and Pharmacokinetics of Cariprazine in Pediatric Patients With ASD. J Am Acad Child Adolesc Psychiatry 2022; 61: S240. | It is an open label, multi-dose study without any placebo comparison. |

6) Study characteristics

| Study | Study type | No. of participants (N), age and gender | Interventions (N) | Dose | Trial duration | Sponsorship/ funding | Methods used for ASD diagnosis | Presence of ID and severity? | Primary & Secondary outcome scales | Findings |
|---|---|---|---|---|---|---|---|---|---|---|
| Aman et al., 2009 | Parallel design RCT of a combination treatment of risperidone + parent training (COMB) vs risperidone alone (MED) with blinded evaluation | 124 children, 95 completed.<br>105 boys (85% of sample) and 19 girls (15%)<br><br>Age 4 - 13 years<br><br>Mean age in Med group: 7.50<br><br>Mean age in Comb group: 7.38 | Comb group: Parent training (PT) and Med (N = 75)<br><br>Med [Ris] (N= 49) | 0.5 to 3.5 mg/day for ris; (switch to aripiprazole if ineffective)<br><br>COMB: average 10.9 parental training sessions | 24 weeks | Funded by National Institute of Mental Health by the following RUPP grant | DSM-IV-TR clinical corroborated by the ADI-R | Yes, both groups had some participants with mild to moderate ID<br><br>Med: mild (18.4%), moderate (34.7%) borderline ID (24.5%), average ID (22.5%)<br><br>Comb: mild (19.2%), moderate (17.8%) borderline ID (24.7%), average ID (38.4%) | HSQ<br><br>ABC, including ABC-I<br><br>CYBOCS-PDD<br><br>CGI-I | COMB was superior to MED on HSQ score (p=0.006).<br><br>Groups did not differ on CGI-I scores at the endpoint. Compared with MED, COMB group showed significant reductions (improvement) in ABC-I (p=0.01), Stereotypic Behavior (p=0.04), and Hyperactivity / Noncompliance subscales (p=0.04).<br><br>Both groups had significant gains in height and substantial gains in weight and BMI. As compared by analysis of covariance, the differences between groups were not significant in weight, height, or BMI on percentile-normed growth lines. |

| | | | | | | | | | | |
|---|---|---|---|---|---|---|---|---|---|---|
| DeVane et al., 2019 | Randomised, double-blind, parallel-group study | 61 children and adolescents, 51 completed<br><br>Aged 6–17 years<br><br>Arip Group<br>Median age: 8.5<br>Age range: 6.0–15.1<br>Boys: 25 (81%)<br><br>Ris Group<br>Median age: 8.3<br>Boys: 23 (77%)<br>Age range: 6.3–17.5 | Arip (N = 31)<br><br>Ris (N= 30) | Arip: 2 to 15 mg/day<br><br>Ris: 0.5 to 2.5 mg/day and for those weighing more than 45 kg, the max dose was 3.0 mg/day | 10 weeks (Optional 3 month extensio n phase) | Funded by the National Institute of Child Health and Human Development, National Institutes of Health. | ASD using DSM-IV corroborate d with the ADR-R criteria and ADOS. | Intelligence of participants was measured but not reported. | The ABC-I<br><br>Children's Sleep Habits Questionnaire<br><br>CGI-S<br>CGI-I | Participants in both groups showed a significant improvement on the ABC-I subscale after one week and continued for the remaining nine weeks.<br><br>Improvement was greatest in the risperidone group at every assessment period and was statistically significantly better than that in the aripiprazole group at weeks 3 and 6 ($p<0.05$).<br><br>No dose-limiting adverse events occurred during the dose-titration period. Mean weight gain in the aripiprazole group was significantly less than that in the risperidone group at week 4 ($p=0.033$) and week 10 ($p<0.001$). |
| Ghanizad eh et al., 2013 | Randomised Double Blind Clinical Trial | 59 children and adolescents, 50 completed.<br><br>Age range: 4-18 years old | Arip (N= 29)<br><br>Ris (N=30) | Arip dose: Maximum dose < 40 kg: up to 10 mg/day and | 2 months | Supported by a grant (No: 3135) from Shiraz University of Medical | DSM-IV-TR and ADI-R. | Not reported | ABC scale<br>CGI-I | The risperidone and the aripiprazole groups both showed a statistically significant improvement in all ABC subscales scores at |

| | | | | | | | | | | |
|---|---|---|---|---|---|---|---|---|---|---|
| | | Arip group<br>Mean age: 9.6 (SD = 3.3)<br>Boys = 25 (86.2%)<br><br>Ris Group<br>Mean age: 9.5 (SD = 4.6)<br>Boys = 23 (76.7%) | | >40 kg: 15 mg/day for<br><br>Ris dose<br>Maximum dose < 40 kg: up to 2 mg, and > 40 kg: up to 3 mg/day | | Sciences to Professor Ahmad Ghanizadeh | | | | follow-up. There was no significant intergroup difference for any ABC subscale.<br><br>The rates of adverse effects were not significantly different between the two groups. |
| Hollander et al., 2006 | Double, blind, randomise, controlled trial | 11 children, 8 completed.<br><br>Age range: 6-14 years old.<br><br>A mean age of 9 years.<br><br>9 male and 2 female.<br><br>Olanzapine group<br>Mean age (SD): 9.25 (2.9)<br>Age range: 6.0–14.8<br>Boys: 6<br><br>Placebo<br>Mean age (SD): 8.9 (2.1)<br>Age range: 6.1–11.0 | Olanzapine (N = 6)<br><br>Placebo (N = 5) | Dosages in the olanzapine group ranged from 7.5 mg/day to 12.5 mg/day, with a mean dose of 10 ± 2.04 mg/day. | 8 weeks | This study was supported by an investigator-initiated research grant from Lilly Research Laboratories. | DSM-IV, ADI-R and ADOS | Yes, presence of ID.<br><br>Olanzapine<br>Normal: 2<br>Mild: 2<br>Severe: 2<br><br>Placebo<br>Normal: 2<br>Mild: 3 | CGI-I<br><br>CY-BOCS<br><br>OAS-M | Olanzapine was superior to the placebo on the mean CGI-I score with a significant linear trend x group interaction (p=0.012). 50% on olanzapine versus 20% on placebo were responders.<br><br>No significant change (linear trend x condition interaction) on the C-YBOCS (p=0.777), the OAS-M irritability measure (p = 0.325), or the OAS-M aggression measure (p=0.671).<br><br>Participants in the olanzapine group gained more weight than participants in the control group. |

| | | | | | | | | | | |
|---|---|---|---|---|---|---|---|---|---|---|
| | | Boys: 3<br>Girls: 2 | | | | | | | | |
| Ichikawa et al., 2016 | A Randomized, Double-blind, Placebo-controlled Study | 92 children and adolescents, 89 completed.<br>Mean age: 10.1 (3.2)<br>Male: 75<br><br>Placebo Group<br>Male : 36<br>Mean age: 9.9 (3.1)<br><br>Arip Group<br>Male: 39<br>Mean age: 10.3 (3.3) | Placebo (N = 45)<br><br>Arip (N = 47) | Arip:<br>A mean (±SD) daily dose of 8.2 ± 4.9 mg at endpoint.<br>Maximum dose 15mg/day | 8 weeks | This study was funded by Otsuka Pharmaceutical Co, Ltd (Tokyo, Japan)<br><br>Some of the authors (corresponding author) are part of the Otsuka Pharmaceutical Co, Ltd (Not main author) | DSM-IV-TR criteria and the pervasive developmental disorders autism society Japan rating scale (PARS) | Yes N= 58 (63.0%) patients with intellectual disability were included in the trial but profound ID were excluded.<br><br>Placebo<br>Mild 16 (35.6)<br>Moderate 7 (15.6)<br>Severe 6 (13.3)<br><br>Arip<br>Mild 16 (34.0)<br>Moderate 7 (14.9)<br>Severe 6 (12.8) | ABC-J irritability<br><br>CGI-I and CGI-S<br><br>CY-BOCS<br><br>CGAS | Aripiprazole was significantly more efficacious than placebo at treating irritability, as measured by the caregiver-rated ABC-I subscale score from week 3 through week 8 (p=0.044).<br><br>Aripiprazole produced significant improvements over the placebo on CGI-I score, mean ABC-hyperactivity subscale score, mean CGI-S score and CAS score. No significant difference between the groups in the mean ABC-stereotypy score.<br><br>Significantly more responders were in the aripiprazole group than in the placebo group (p=0.033).<br><br>No serious adverse event was reported in the aripiprazole group. |

| | | | | | | | | | | |
|---|---|---|---|---|---|---|---|---|---|---|
| Kent et al., 2013 | A Double-Blind, Placebo-Controlled Study | 96 children, 77 completed<br><br>Mean age of 9 (3.1) years old, 77 % (N= 74) were <12 years of age.<br><br>Boys: 84 (88%)<br><br>Low dose<br>Mean age: 10 (3.4)<br>Boys: 25 (83%)<br><br>High dose<br>Mean age: 9 (3.1)<br>Boys: 28 (90%)<br><br>Placebo<br>Mean age:<br>9 (2.6)<br>Boys: 31 (89%) | Two fixed-dose and placebo.<br><br>Low dose Ris (N= 30)<br><br>High dose of Ris (N= 31)<br><br>Placebo (N = 35) | Risperidone low dose: fixed-dose 0.125 mg/day (<45kg) or 0.175 mg/day (>45kg)<br><br>Risperidone high dose: fixed-dose 1.25 mg/day (<45kg) or 1.75 mg/day (>45kg) | 6 weeks<br><br>(+6 months open label) | This study was funded by Johnson & Johnson Pharmaceutical Research & Development, LLC | DSM-IV and ADI-R | Not reported but the median mental age across treatment groups was 5.5 years. | ABC including ABC-I<br><br>CGI-S score<br><br>CYBOCS<br><br>CGI-I | Mean baseline to endpoint change in ABC-I was significantly greater in the high-dose ($p<0.001$) but not the low-dose ($p=0.164$) group versus placebo.<br><br>CGI-S and C-YBOCS scores improved significantly only in the high-dose group.<br><br>Somnolence, sedation and increased appetite occurred more frequently in high- versus low-dose groups. |
| Kouhbanani et al., 2021 | A randomised controlled trial | 45 Children, 43 completed.<br><br>Age range: 6-12 years old.<br><br>Ris<br>Mean age: 8.44 (1.94)<br>Age range: 6–11<br>Boys: 11 (73%)<br><br>VR+Ris5 | ris (N= 15)<br><br>VR + ris (N= 15),<br>90 sessions of training<br><br>Passive control group (N= 15) | Risperidone: maximum dose 1.75mg/day (<20kg) or 2.25mg/day (20-45kg) or 3.5mg/day (>45kg) | Three months | Not reported | DSM-V, ADI-R and CARS | All participants had low to moderate IQ. | The CARS-II<br><br>VABS | Risperidone + VR group showed a statistically significant improvement in social skills ($p<0.001$) and behavioral symptoms ($p<0.001$) at follow-up compared with the risperidone-only group. |

| | | | | | | | | | | |
|---|---|---|---|---|---|---|---|---|---|---|
| | | Mean age: 8.59 (2.12)<br>Age range: 6–12<br>Boys: 11 (75%)<br><br>Control<br>Mean age: 8.40 (2.01)<br>Age range: 6–12<br>Boys: 10 (69%) | | | | | | | | |
| Loebel et al., 2016 | Randomized, double-blind, fixed-dose, placebo-controlled study | 150 children and adolescents, 128 completed.<br>Age range: 6–17 years.<br><br>Lur low dose group:<br>Mean age (SD): 10.5 (3)<br>Boys: 38 (79.2%)<br><br>Lur high dose group:<br>Mean age (SD): 10.5 (3)<br>Boys: 43 (84.3%)<br><br>Placebo:<br>Mean age (SD): 11 (3)<br>Boys: 40 (81.6%) | Lur low dose of 20 mg/day (N = 50)<br><br>Lur High dose of 60 mg/day (N = 49)<br><br>Placebo (N = 51) | Fixed, once-daily doses of lurasidone (20 or 60 mg/day). | 6 weeks | The original clinical research was sponsored by Sunovion Pharmaceuticals Inc. The sponsor was involved in the design, collection, and analysis of the data. | DSM-IV-TR and ADI-R | Not reported.<br><br>However, participants were excluded if a confirmed genetic disorder associated with cognitive and/or behavioral disturbance or if participant has profound intellectual disability. | ABC-I<br>CGI-I<br>CYBOCS<br>CGSQ | The least squares (LS) mean improvement from baseline to week 6 in the ABC-I was not significantly different for lurasidone 20 mg/day and lurasidone 60 mg/day versus placebo.<br><br>CGI-I scores showed significantly greater LS mean improvement at week 6 for lurasidone 20 mg/day versus placebo ($p = 0.035$) but not for lurasidone 60 mg/day.<br><br>Discontinuation rates due to adverse events were no higher in lurasidone groups. Adverse events with an incidence >10% (lurasidone combined |

| | | | | | | | | | | |
|---|---|---|---|---|---|---|---|---|---|---|
| | | | | | | | | | | dose, placebo) included vomiting and somnolence.<br><br>Modest changes were observed in weight and selected metabolic parameters. |
| Luby et al., 2006 | A randomized, double blind, placebo-controlled | 24 children (one excluded as did not meet criteria)<br><br>Age range: 2.5 and 6.0 years old. Male: 17<br><br>Ris Group<br>Age in months: Mean (SD): 49.0 (10.9)<br>Boys: 9<br><br>Placebo Group<br>Age in months: Mean (SD): 48.1 (13.2)<br>Boys: 8 | Ris (N=11)<br><br>Placebo (N=12) | Ris dose range: 0.5–1.5 mg | 6 months | This study was funded by Janssen Pharmaceutica as an investigator initiated project to Dr. Luby. | DSM-IV | Measured but not reported | VABS<br>CBCL<br>PLS-3<br>CARS<br>GARS | Controlling for baseline intergroup differences, pre-schoolers on risperidone demonstrated greater improvements in autism severity.<br><br>The change in autism severity scores from baseline to 6-month follow-up for the risperidone group was 8% compared to 3% for the placebo group. Notably, both groups significantly improved over the 6-month treatment period.<br><br>Preschool children tolerated low-dose risperidone well, with no serious adverse effects observed over a 6-month treatment period. Weight gain and hypersalivation were |

| | | | | | | | | | | |
|---|---|---|---|---|---|---|---|---|---|---|
| | | | | | | | | | | the most common side effects reported, and hyperprolactinemia without lactation or related signs was observed. |
| Mahajan et al., 2022 | A randomized, open-label trial | 49 children and adolescents, 40 completed.<br><br>Age range: 6 and 16 years<br><br>Mean age (SD): 8.05 (2.31) years. Male: 35<br><br>MPH<br>Mean age (SD): 7.25 (2.0)<br>Boys: 17<br><br>Ris<br>Mean age (SD): 8.85 (2.6)<br>Boys: 18 | MPH (N= 25)<br><br>Ris (N = 24) | Mean MPH= 19.13 ± 7.0 mg/day.<br><br>Ris= 0.5 mg for < than 20 kg, and 1 mg/day > 20 kg | 8 weeks | Not funded | DSM-5 | 20/ 40 (50%) had mild intellectual disability. | CARS-II<br>ISAA<br>N-CBRF | There was an overall significant improvement in the final scores of ISAA and CARS-2 for both Ris and MPH groups, with no group showing significantly higher improvement than the other.<br><br>Only the activity level item within the CARS-2 was statistically significant for both groups, contributing towards the significant improvement in the total score. In the ISAA scale, both groups showed improvement in the behavior patterns and cognitive component subscales, but only the Ris group showed an improvement in the emotional |

| | | | | | | | | | | |
|---|---|---|---|---|---|---|---|---|---|---|
| | | | | | | | | | | responsiveness subscale.<br><br>Side effects were greater in the MPH (N=11) group compared to the RSN group (N=8). |
| Marcus et al., 2009 | A double-blind, randomized, placebo-controlled, parallel-group study. | 218 children, and adolescents, 178 completed.<br>Aged 6-17 years old<br>Mean age: 9.7 years old<br><br>Arip<br>low dose group<br>Mean age (SD): 9.0 (2.8)<br>Boys: 47 (88.7%)<br><br>Medium dose group<br>Mean age (SD): 10.0 (3.2)<br>Boys: 50 (84.7%)<br><br>High dose group<br>Mean age<br>(SD): 9.5 (3.1)<br>Boys: 50 (92.65%)<br><br>Placebo | Arip<br>Low dose 5 mg/day (N=53)<br><br>Medium dose 10 mg/day (N=59)<br><br>High dose 15 mg/day (N=54)<br><br>Placebo (N=52) | Fixed doses of 5/10/15 mg/day. | 8 weeks | This study was supported by Bristol-Myers Squibb (Princeton, NJ) and Otsuka Pharmaceutical Co., Ltd. (Tokyo, Japan).. | DSM-IV-TR<br>ADI-R | Not reported | ABC-I<br>CGI-I<br>CGI-S<br>CYBOCS<br>PedsQL<br>CGSQ | At week 8, all aripiprazole doses produced significantly greater improvement than placebo in mean ABC-I subscale scores (p<0.05).<br><br>All aripiprazole doses demonstrated significantly greater improvements in mean CGI-I score than placebo at week 8.<br><br>The most common adverse event leading to discontinuation was sedation. There were two serious adverse events: presyncope (5 mg/day) and aggression (10 mg/day). There was significant weight gain in the aripiprazole groups (p<0.05 versus placebo). |

| | | | | | | | | | | |
|---|---|---|---|---|---|---|---|---|---|---|
| | | Mean age (SD): 10.2 (3.1)<br>Boys: 48 (92.3%) | | | | | | | | |
| Martsenkovska, 2014 | double-blind, randomized controlled study | 86 children<br><br>Age range: 3 - 6 years old.<br><br>Ris Group<br>Mean Age: 4.95 (SD=0.36)<br>Boys = 31; Girls =12<br><br>Divalproex sodium Group<br>Mean Age: 4.87 (SD=0.41)<br>Boys =27 ; Girls= 16 | Ris (N= 43)<br><br>Divalproex sodium (N= 43) | Mean daily dosage of Ris was 0.005-0.01 mg/kg.<br><br>Mean daily dosage of divalproex sodium was titrated up to effect and/or valproate level between 50 and 100 pg/ml. | 16 weeks | Not reported | DSM-IV and confirmed by the ADI-R and the ADOS | Not reported<br>But<br>children with severe mental retardation in whom a definitive diagnosis of autism could not be made were also excluded. | CGI-I<br>OAS-M<br>ABC-I | Risperidone is significantly better than Divalproex in improving irritability measured by CGI-I (p=0.002, d=1.46); OAS-M aggression against objects (p=0.005); ABC-I (teacher rating) (p=0.05). |
| McCracken, 2002 | Multisite, randomised, double-blind trial | 101 children, 80 completed.<br>82 boys and 19 girls<br>Mean [±SD] age: 8.8±2.7 years old<br>Agre range: 5 to 17 years old.<br><br>Ris Group<br>Boys: 39 | Ris (N = 49)<br><br>Placebo (N=52) | Dose range, 0.5 to 3.5 mg per day | 8 weeks | Supported by contracts from the National Institute of Mental Health and a grant from the Korczak Foundation. | DSM-IV<br>ADI-R | Required to have a mental age of at least 18 months.<br><br>Average or above-average IQ:<br>Ris – 3<br>Placebo – 2<br><br>Borderline IQ:<br>Ris – 8 | ABC (including ABC-I)<br>CGI-I | After eight weeks of treatment, the risperidone group had a significant decrease (improvement) in the mean ABC-I score compared with the placebo group (p<0.001). |

| | | | | | | | | | | |
|---|---|---|---|---|---|---|---|---|---|---|
| | | Placebo Group<br>Boys: 43 | | | | | | Placebo – 4<br><br>Mild or moderate retardation:<br>Ris – 20<br>Placebo – 23<br><br>Severe retardation:<br>Ris – 15<br>Placebo – 16 | | The rate of a positive response (at least a 25% improvement in the score on the ABC-I and a rating of much improved or very much improved on the CGI-I scale) was significantly higher in the risperidone group (p<0.001).<br><br>Risperidone therapy was associated with higher weight gain compared with placebo (p<0.001). Increased appetite, fatigue, drowsiness, dizziness, and drooling were more common in the risperidone group than in the placebo group (p<0.05 for each comparison). |
| McDougle 1998 | A double blind, placebo-controlled study. | 31 Adults, 24 completed.<br>18-43 Years old<br><br>Mean age (SD): 28.1 (7.3)<br><br>Ris<br>Mean age (SD): 26 (6.7)<br>Men: 13 | Ris (N = 15)<br><br>Placebo (N= 16) | Mean daily dose of Ris (SD): 2.9 (1.4)<br><br>Mean daily dose of placebo (SD): 3.9 (1.5) | 12 weeks | This study was supported in part by various grants including from the Public Health Service Young Investigator Award and an Independent | DSM-IV<br>ADI-R<br>ADOS | Mean IQ (SD): 54.6 (23.9)<br><br>Mean IQ of Ris:<br>55 (26.8)<br>Mean IQ of placebo: 52.9 (22.1)<br><br>Average IQ:<br>Ris -3<br>Placebo-1 | Y-BOCS<br>SIB-Q<br>RF-RLRS<br>CGI-I<br>VAS | 57% of the patients treated with risperidone were categorized as responders compared with none of the placebo group (p<0.002).<br><br>Risperidone was superior to placebo in |

| | | | | | | | | | | |
|---|---|---|---|---|---|---|---|---|---|---|
| | | Age range: 18-38 years old<br><br>Placebo<br>Mean age (SD) : 29.7 (7.8)<br>Men: 9<br>Age range: 18- 43 years old. | | | | Investigator Award; Theodore and Vada Stanley Foundation Research Awards Program; State of Connecticut, Department of Mental Health and Addiction Services; RUPP; and National Institute of Mental Health. | | Borderline IQ:<br>Ris -1<br>Placebo- 2<br><br>Mild IQ<br>Ris -4<br>Placebo- 6<br><br>Moderate IQ<br>Ris -3<br>Placebo-4<br><br>Severe IQ<br>Ris -4<br>Placebo-3 | | reducing repetitive behavior (p<0.001), aggression (p<0.001), anxiety or nervousness (p<0.02), depression (p<0.03), irritability (p<0.01), and overall behavioral symptoms of autism (p<0.02). Objective, measurable changes in social behavior and language did not occur.<br><br>Other than mild, transient sedation, risperidone was well tolerated, with no evidence of extrapyramidal, cardiac or seizure related symptoms. |
| Miral et al., 2008 | A randomize, controlled, double-blind study | 30 children and adolescents, 28 completed.<br>Age range: 8 – 18 years old.<br>24 were boys and 6 were girls.<br><br>Ris Group<br>Mean age (SD): 10.0 (±2.7)<br>Mean age: 7–15<br>Boys: 11 | Ris (N=15)<br><br>Hal (N=15) | Starting dosage of 0.01 mg/kg/day and the dosage was increased to 0.04 mg/kg/day until the end of the first 2 weeks. If tolerated, then it was | 12 weeks | This research was supported in part by Janssen and Cilag Drug Company. | DSM-IV | Not reported. | RF-RLRS<br>ABC<br>CGI-S<br>CGI-I | The reduction from baseline in RF-RLRS, sensory-motor (subscale I) and language (subscale V) scores were significant in the risperidone group (p<0.05).<br><br>Compared with haloperidol, the risperidone group showed a significantly greater reduction |

| | | | | | | | | | | |
|---|---|---|---|---|---|---|---|---|---|---|
| | | Hal Group<br>Mean age (SD): 10.9 (±2.9)<br>Mean age: 7-17<br>Boys:13 | | increased to a maximum dosage of 0.08 mg/kg/day. | | | | | | (improvement) in the ABC and DSM-IV PDD scale scores ($p<0.05$ and $p<0.01$).<br><br>Sensory motor behaviors (subscale I) and language at the end of the 12th week, and RF-RLRS sensory motor and language subscale scores improved significantly in the risperidone than the haloperidol group ($p<0.05$).<br><br>There was a greater increase of serum prolactin level in the risperidone group, while liver function tests showed a poorer result in the haloperidol group. |
| Nagaraj et al., 2006 | Randomized, Placebo-Controlled, Double-Blind Study | 40 children, 39 completed.<br>Age range: 2 to 9 years old<br><br>Ris Group<br>Age (in months) (SD): 57.95 ± 20.84<br>Boys: 16 (84.2%) | Ris (N=20)<br><br>Placebo (N = 20) | dose of 1 mg/day<br><br>Starting dose: 0.5 mg daily orally for the first 2 weeks | 6 months | This study was entirely supported by the Department of Pediatrics and the i n s t i t u t e 's internal finances. | DSM-IV | Children with severe mental retardation were excluded.<br><br>Borderline IQ:<br>Ris- 9 (47.4%)<br>Placebo- 8 (40%)<br><br>Mild retardation: | CARS<br>CGAS<br>VSMS<br>Global Impression of Parents (Parent questionnaire developed by the authors) | In the risperidone group, significantly more children showed improvement in the total CARS score and CGAS score compared with the placebo group ($p<0.001$ and $p=0.035$, respectively). |

| | | | | | | | | | | |
|---|---|---|---|---|---|---|---|---|---|---|
| | | Placebo Group<br>Age (in months) (SD): 63.0 ± 20.12<br>Boys: 18 (90%) | | | | | | Ris- 6 (31.6%)<br>Placebo- 5 (25%)<br><br>Moderate retardation:<br>Ris- 4 (21.1%)<br>Placebo- 7 (35%) | | Risperidone significantly improved functioning in the domains of social responsiveness (p=0.014) and nonverbal communication (p=0.008) and decreased symptoms of hyperactivity (p=0.002) and aggression and irritability (p=0.016).<br><br>Risperidone was associated with increased appetite and mild weight gain, mild sedation in 20%, and transient dyskinesias in three children. |
| NCT00198107 | Randomized, Placebo-Controlled, Quadruple blind study | 81 children and adolescents, 72 completed.<br>Mean age (SD): 9.2 (3.2)<br>Boys: 70<br><br>Arip Group<br>Mean age (SD): 9.0 (2.9)<br>Boys: 36<br><br>Placebo Group<br>Mean age (SD): 9.4 (3.5) | Arip (N = 40)<br><br>Placebo (N= 41) | For participants weighing less than 50 kg, the maximum dose will be 10 mg per day. For more than 50 kg, maximum dose will be 15 mg. | 8 weeks | Supported by NIH grants. | DSM-IV | Not reported. | ABC and ABC-I<br>CGI-I<br>CY-BOCS<br>VABS<br>ADOS<br>SRS | Significant reduction in the ABC-I score in the aripiprazole group (post-intervention score: 18.6, 95% CI 15.8-21.4) compared with the placebo group (post-intervention score: 25.5, 95% CI 22.8-28.3) (p=0.0006).<br><br>Improvement in CGI-I score with aripiprazole (odds ratio 0.5, 95% CI 0.31-0.79) compared |

| | | | | | | | | | | |
|---|---|---|---|---|---|---|---|---|---|---|
| | | Boys: 34 | | D - cycloserine: range of 25 to 200 mg daily | | | | | | with placebo (odds ratio 0.1, 95% CI 0.05-0.23). |
| NCT00468130 | A randomised, Placebo-Controlled, Quadruple blind study | 13 children, 9 completed.<br>Mean age (SD): 12.4 (2)<br><br>Arip Group<br>Mean age (SD): 12.3 (2)<br>Boys: 6<br><br>Placebo Group<br>Mean age (SD): 12.6 (2)<br>Boys: 5 | Arip (N=7)<br><br>Placebo (N= 6) | <40 kg: started on 2.5mg per day in 1st week and and increased to 5 mg at week 2. Max dose is 10 mg.<br><br>>40 kg: start at 5 mg and increase to 10 mg at week 2 until they reach a maximum of 20 mg at week 4. | 8-week | Not reported | DSM-IV<br>ADI-R | Not reported | CGI-AD<br>ABC-I | CGI-I: Aripiprazole: baseline (3.83 ± 0.41) vs. FU (2.67 ± 1.21); placebo: baseline (4.25 ± 1.25) vs. FU (4.25 ± 1.5) (p=0.06).<br><br>ABC-I: Aripiprazole: baseline (15.67 ± 11.65) vs. FU (6.83 ± 6.7); placebo: baseline (8 ± 4.58) vs. FU (8.67 ± 10.69) (p=0.725).<br><br>Adverse event (worsening of depression): Aripiprazole: (1/7) (14.29%); placebo: 0% |
| NCT01624675 | A Double-blind, Placebo-controlled Study, Followed by an Open-label | 39 children, 29 completed<br><br><br>Ris Group<br>Median age: 8.0 years old.<br>Boys: 16 (76.2%) | Ris (N=21)<br><br>Placebo (N= 18) | <20 kg: The starting dose of 0.25 mg/day increased to 0.5 mg/day on day 4.<br>>20 kg: starting dose | 8 weeks | Funder not reported but sponsor is Janssen Pharmaceutical K.K. | DSM-IV-TR | IQ measured but not reported.<br><br>Include only patients with mental age or development age of >18 months and IQ of >35. | ABC-J and ABC-J Irritability<br>PSQ<br>CGI-C<br>CGI-S<br>CGAS | Statistically significant improvement in ABC-I score for the risperidone group compared with the placebo group (p=0.003). |

| | | | | | | | | | | |
|---|---|---|---|---|---|---|---|---|---|---|
| | Extension Study | Placebo Group<br>Median age: 7.0 years old<br>Boys: 14 (77.8%) | | of 0.5 mg/day and increased to 1.0 mg/day on day 4. >45 kg: max dose of 3.0 mg. | | | | | | No statistically significant between-group difference was found in the change of the lethargy/social withdrawal subscale score from baseline to the endpoint (ANCOVA, p=0.6409), while the other three subscale scores showed statistically significant improvements in the risperidone group compared with the placebo group (p=0.0353 on the stereotypic behavior, p=0.0042 on the hyperactivity/noncompliance, and p=0.0364 on the Inappropriate Speech)<br><br>No statistically significant difference between CGI-C and CGI-S.<br><br>Statistically significant improvement for CGAS in the risperidone group compared with the placebo group (p=0.0045). |

| | | | | | | | | | | |
|---|---|---|---|---|---|---|---|---|---|---|
| Nikvarz et al., 2016 | A randomized, open-label trial | 34 children, 30 completed<br>Age range 4–17 years old<br>Mean age (SD): 6.7±3.2 years old<br>Boys: 23 (76.7%)<br><br>Ris Group<br>Mean age (SD): 6.56 (3.51)<br>Boys: 10 (66.7%)<br><br>Mem Group<br>Mean age (SD): 6.83 (2.98)<br>Boys: 13 (86.7%) | Ris (N = 16)<br><br>Mem (N = 18) | Ris<br>Max dose: 3mg/day<br><br>Mem:<br>Max dose: 20mg/day. | 8 Weeks | This study was supported by Tehran University of Medical Sciences (TUMS) (Grant number: 91-01-33-16991 | DSM-IV-TR | Not reported | ABC<br>CARS<br>CGI-I and CGI-S | Both risperidone and memantine reduced the scores of 4 ABC subscales as well as the 10-item and the total score of CARS significantly at follow-up. However, there was no statistically significant intergroup difference in any of these scores.<br><br>Relatively, a larger number of participants on risperidone showed “very much improvement” when assessed by the CGI-I scale compared with those on memantine. |
| Owen et al., 2009 | A double-blind, randomized, placebo-controlled, parallel-group study | 98 children and adolescents, 75 completed.<br>Age range 6 –17 years.<br>Mean age: 9.3 years old.<br><br>Arip Group<br>Age, mean (SD): 9.7 (3.2)<br>Boys: 42 (89.4%)<br><br>Placebo Group | Arip (N= 47)<br><br>Placebo (N= 51) | Arip = starting dose 2 mg/day, with a target dosage of 5, 10, or 15 mg/day (maximum dosage: 15 mg/day) | 8 weeks | This study was supported by Bristol Myers Squibb (Princeton, NJ) and Otsuka Pharmaceutical Co, Ltd (Tokyo, Japan). | DSM-IV-TR corroborated by ADI-R. | Not reported. | CGI-I<br>ABC and ABC-I<br>CGSQ<br>PedsQL<br>CYBOCS | Mean improvement in ABC-I score was significantly greater in the aripiprazole than the placebo group from week 1 through week 8 (p<0.001).<br><br>Aripiprazole demonstrated significantly greater global improvements than placebo, as assessed by the mean CGI-I score from week 1 |

| | | | | | | | | | | |
|---|---|---|---|---|---|---|---|---|---|---|
| | | Age, mean (SD): 8.8 (2.6)<br>Boys: 44 (86.3%) | | | | | | | | through week 8 (p<0.001). However, clinically significant residual symptoms may still persist for some patients.<br><br>Discontinuation rates as a result of adverse events and EPS-related adverse event rates were slightly higher for the aripiprazole group. No serious adverse events were reported. Aripiprazole treatment was associated with significantly greater mean weight change compared with placebo at the endpoint (p=0.005). |
| Shea et al., 2004 | A randomized, double-blind, placebo-controlled trial | 80 children, 72 completed.<br>Age range: 5 to 12 years<br>Mean Age: 7.5 years<br><br>Ris Group<br>Mean age, SE: 7.6 (2.3)<br>Median age (range): 7.0 (5–12)<br>Boys: 29 (72.5) | Ris (N = 41)<br><br>Placebo (N = 39) | 0.01– 0.06 mg/kg/day<br><br>Ris mean daily dose 1.48 mg, and the mean dosage was 0.05 mg/kg/day | 8 weeks | This trial was supported by Janssen-Ortho Inc, Canada, and Johnson & Johnson Pharmaceutical Research and Development. | DSM-IV<br>CARS | Only tested IQ on 31/40 in Ris and 35/39 in the placebo group.<br><br>Normal<br>Ris- 3<br>Placebo – 11<br><br>Borderline<br>Ris- 6<br>Placebo – 4<br><br>Mild | ABC<br>CGI-I, CGI-S, CGI-C<br>N-CBRF<br>VAS | Participants taking risperidone showed a significantly greater mean decrease in the ABC-I score compared with those taking a placebo (p<0.001).<br><br>The risperidone-treated group also showed a significant decrease on the other four subscales of the ABC (p<0.05) and on the |

| | | | | | | | | | | |
|---|---|---|---|---|---|---|---|---|---|---|
| | | Placebo Group<br>Mean age, SE: 7.3 (2.3)<br>Median age (range): 7.0 (5–12)<br>Boys: 32 (82.1) | | | | | | Ris- 12<br>Placebo – 8<br><br>Moderate<br>Ris- 10<br>Placebo – 12 | | conduct problem, insecure/anxious, hyperactive, and overly sensitive subscales of the N-CBRF (parent version); and on VAS of the most troublesome symptom.<br><br>More risperidone-treated participants showed global improvement in their condition compared with the placebo group.<br><br>Risperidone-treated participants showed more somnolence and statistically significantly greater increases in weight, pulse rate, and systolic blood pressure. Extrapyramidal symptom scores were comparable between groups. |

ABC= Aberrant Behavior Checklist; ABC-C rating scale= Aberrant Behavior Checklist-Community; ABC-I = Aberrant Behaviour Checklist Irritability subscale; ABC-J = Aberrant Behaviour Checklist Japanese Version; Arip= Aripiprazole; ADI-R= Autism Diagnostic Interview-Revised; ADOS= the Autism Diagnostic Observation Schedule; CARS = Childhood Autism Rating Scale; CARS-II = The Children Autism Rating Scale II Test; CBCL = The Childhood Behavior Checklist; CGAS = Children's Global Assessment Scale; CGI-AD = Clinical Global Impression Improvement, modified for autistic disorder; CGI-C = Clinical global impression-change; CGI-S = Clinical Global Impression–Severity; CGI-I = Clinical Global Impression–Improvement; CGSQ= Caregiver Strain Questionnaire; CY- BOCS = Children's Yale–Brown Obsessive Compulsive Scale; CYBOCS-PDD = Children's Yale-Brown Obsessive Compulsive Scale- PDD version; COMB= Combination treatment of Risperidone and parent training; DSM-IV = Diagnostic and Statistical Manual of Mental Disorders, 4th edition; DSM-IV-TR = The Diagnostic and Statistical Manual of Mental

Disorders, fourth edition, text revision; DSM-5= the Diagnostic and Statistical Manual of Mental Disorders, 5th edition; GARS= Gilliam Autism Rating Scale; Hal= Haliperidol; HSQ= Home Situations Questionnaire; ISAA= Indian Scale for Assessment of Autism; Lur = lurasidone; Mem = Memantine; N-CBRF= the Nisonger Child Behavior Rating Form; FOAS-M = Overt Aggression Scale–Modified; PedsQL= Pediatric Quality of Life Inventory; PDD = Pervasive developmental disorders; PLS-3 = Preschool Language Scale, Third Edition; PSQ = Parent satisfaction questionnaire; RF-RLRS: Ritvo–Freeman Real Life Rating Scale; RIS = Risperidone; RUPP = Research Unit on Pediatric Psychopharmacology; SIB-Q= Self-injurious Behaviour Questionnaire; SRS= Social Reciprocity Scale; VABS = Vineland Adaptive Behavior Scales, Interview Edition; VAS= Visual Analog Scale; VSMS=Vineland Social Maturity Scale; Y-BOCS = The Yale–Brown Obsessive–Compulsive Scale

7) Risk of bias results

| Study name | Intervention 1 | Intervention 2 | Bias arising from the randomisation process | Risk of bias due to deviations from the intended interventions | Risk of bias due to missing outcome data | Risk of bias in measurement of the outcome | Risk of bias in selection of the reported result | Overall risk of bias |
|---|---|---|---|---|---|---|---|---|
| Aman et al., 2009 | Ris | Ris + PT | Some concerns | Low risk | Low risk | Low risk | Some concerns | Some concerns |
| DeVane et al., 2019 | Ris | Arip | Low risk | Some concerns | Some concerns | Low risk | Low risk | Some concerns |
| Ghanizadeh et al., 2013 | Ris | Arip | Some concerns | Some concerns | Low risk | Low risk | Low risk | Some concerns |
| Hollander et al., 2006 | Olanz | Placebo | Some concerns | Low risk | Low risk | Low risk | Some concerns | Some concerns |
| Ichikawaet al., 2016 | Arip | Placebo | Low risk | Low risk | Low risk | Low risk | Low risk | Low risk |
| Kent et al., 2013 | Placebo | Ris – low dose, and Ris - high dose | Low risk | Low risk | Low risk | Low risk | Low risk | Low risk |
| Kouhbanani et al., 2021 | placebo | VR + Ris, and Ris | Some concerns | High risk | Some concerns | Low risk | Low risk | High risk |
| Loebel et al., 2016 | Lur low dose and Lur high dose | Placebo | Low risk | Low risk | Low risk | Low risk | Low risk | Low risk |
| Luby et al., 2006 | Ris | Placebo | High risk | Some concerns | Low risk | Low risk | Low risk | High risk |
| Mahajan et al., 2022 | MPH | Ris | Low risk | Some concerns | Some concerns | Some concerns | Low risk | Some concerns |
| Marcus et al., 2009 | Arip low, medium and high dose | Placebo | Some concerns | Low risk | Some concerns | Low risk | Low risk | Some concerns |
| Martsenkovska, 2014 | Ris | Divalproex sodium | Some concerns | Some concerns | Low risk | Low risk | High risk | High risk |
| McCracken, 2002 | Ris | Placebo | Some concerns | Low risk | Low risk | Low risk | Low risk | Some concerns |
| McDougle 1998 | Ris | Placebo | Some concerns | Low risk | Low risk | Low risk | Low risk | Some concerns |
| Miral et al., 2008 | Ris | Hal | Some concerns | High risk | High risk | High risk | Low risk | High risk |
| Nagaraj et al., 2006 | Ris | Placebo | Low risk | Low risk | Low risk | Low risk | Low risk | Low risk |
| NCT00198107 | Arip | Placebo | Some concerns | Some concerns | Low risk | Low risk | Some concerns | Some concerns |
| NCT00468130 | Arip | Placebo | Some concerns | Some concerns | Low risk | Low risk | Some concerns | Some concerns |

| | | | | | | | | |
|---|---|---|---|---|---|---|---|---|
| NCT01624675 | Ris | Placebo | Some concerns | Low risk | Some concerns | Low risk | Low risk | Some concerns |
| Nikvarz et al., 2016 | Ris | Mem | Some concerns | Low risk | Some concerns | Some concerns | Low risk | Some concerns |
| Owen et al., 2009 | Arip | Placebo | Low risk | Low risk | Low risk | Low risk | Low risk | Low risk |
| Shea et al., 2004 | Ris | Placebo | Some concerns | Low risk | Low risk | Low risk | Low risk | Some concerns |
