## Supplementary material 2 for "A network meta-analysis of randomised controlled trials of antipsychotic medications to assess their comparative efficacy and tolerability in autistic people"

Supplementary material 2: descriptive assessment of transitivity assumption

| Covariate | Ris vs Placebo | Arip vs Placebo | Arip vs Ris | Lur vs Placebo | Ris_PT vs Ris | Mem vs Ris | VPA vs Ris | Hal vs Ris | Olanz vs Placebo | MPH vs Ris |
|---|---|---|---|---|---|---|---|---|---|---|
| ID | N (%) | N (%) | N (%) | N (%) | N (%) | N (%) | N (%) | N (%) | N (%) | N (%) |
| Presence of borderline ID or ID (moderate and below) in some participants | 4 (57.14%) | 1 (20%) | | | 1 (100%) | | | | 1 (100%) | 1 (100%) |
| No presence of ID in all participants | | | | | | | | | | |
| Not reported | 3 (42.86%)<br><br>(Kent, Luby, NCT01624675) | 4 (80%) (Marcus, Owen, NCT00468130, NCT00198107) | 2 (100%) | 1 (100%) | | 1 (100%) | 1 (100%) | 1 (100%) | | |
| Age | N (%) | N (%) | N (%) | N (%) | N (%) | N (%) | N (%) | N (%) | N (%) | N (%) |
| Children and adolescents | 6 (85.71%) | 5 (100%) | 2 (100%) | 1 (100%) | 1 (100%) | 1 (100%) | 1 (100%) | 1 (100%) | 1 (100%) | 1 (100%) |
| Adult | 1 (14.29%) (McDougle) | | | | | | | | | |
| Presence of psychiatric disorders | N (%) | N (%) | N (%) | N (%) | N (%) | N (%) | N (%) | N (%) | N (%) | N (%) |
| Yes | | | | | | | | | | |
| No | 6 (85.71%) | 5 (100%) | | 1 (100%) | 1 (100%) | | | 1 (100%) | 1 (100%) | 1 (100%) |
| Not reported | 1 (14.29%) (Nagaraj) | | 2 (100%) | | | 1 (100%) | 1 (100%) | | | |
| Presence of BtC | N (%) | N (%) | N (%) | N (%) | N (%) | N (%) | N (%) | | N (%) | N (%) |
| Yes | 7 (100%) | 5 (100%) | 2 (100%) | 1 (100%) | 1 (100%) | 1 (100%) | 1 (100%) | 1 (100%) | 1 (100%) | 1 (100%) |
| No | | | | | | | | | | |

Supplementary material 2: descriptive assessment of transitivity assumption

| | | | | | | | | | | |
|---|---|---|---|---|---|---|---|---|---|---|
| Not reported | | | | | | | | | | |
| Novel medication | N (%) | N (%) | N (%) | N (%) | N (%) | N (%) | N (%) | N (%) | N (%) | N (%) |
| Yes | 7 (100%) | 5 (100%) | 2 (100%) | 1 (100%) | 1 (100%) | 1 (100%) | 1 (100%) | 1 (100%) | 1 (100%) | 1 (100%) |
| No | | | | | | | | | | |
| Not reported | | | | | | | | | | |

Covariates per outcomes

| ABC-I | Yes | No | Not reported |
|---|---|---|---|
| ID | 4 | | 10 |
| Psychiatric illness | | 11 | 3 |
| BtC | 14 | | |
| Novel medication | 14 | | |

| ABC-I | Children | Adults | Nor reported |
|---|---|---|---|
| Age | 14 | | |

| CGI-I | Yes | No | Not reported |
|---|---|---|---|
| ID | 2 | | 9 |
| Psychiatric illness | | 8 | 3 |
| BtC | 11 | | |
| Novel medication | 10 | | 1 |

| CGI-I | Children | Adults | Nor reported |
|---|---|---|---|
| Age | 10 | 1 | |

| Overall AE | Yes | No | Not reported |
|---|---|---|---|

Supplementary material 2: descriptive assessment of transitivity assumption

| | | | |
|---|---|---|---|
| ID | 4 | | 8 |
| Psychiatric illness | | 11 | 1 |
| BtC | 12 | | |
| Novel medication | 12 | | |

| Overall AE | Children | Adults | Nor reported |
|---|---|---|---|
| Age | 11 | 1 | |

| Drop out due to AE | Yes | No | Not reported |
|---|---|---|---|
| ID | 6 | | 9 |
| Psychiatric illness | | 14 | 1 |
| BtC | 15 | | |
| Novel medication | 15 | | |

| Drop out due to AE | Children | Adults | Nor reported |
|---|---|---|---|
| Age | 14 | 1 | |

| Sedation | Yes | No | Not reported |
|---|---|---|---|
| ID | 7 | | 9 |
| Psychiatric illness | | 12 | 4 |
| BtC | 16 | | |
| Novel medication | 16 | | |

| Sedation | Children | Adults | Nor reported |
|---|---|---|---|
| Age | 15 | 1 | |

| Weight gain | Yes | No | Not reported |
|---|---|---|---|
| ID | 5 | | 8 |

Supplementary material 2: descriptive assessment of transitivity assumption

| | | | |
|---|---|---|---|
| Psychiatric illness | | 11 | 2 |
| BtC | 13 | | |
| Novel medication | 13 | | |

| Weight gain | Children | Adults | Nor reported |
|---|---|---|---|
| Age | 12 | 1 | |

Studies in each network:

| | |
|---|---|
| Ris vs Placebo | Kent, NCT01624675, Shea 2004, McCracken 2002, Luby et al 2006, McDougle et al., 1998, Nagaraj et al., 2006, |
| Arip vs plaebo | Ichikawa 2017, Marcus 2009, Owen 2019, NCT00468130, NCT00198107 |
| Arip vs Ris | Ghanizadeh 2014, De Vane 2019 |
| Lur vs Placebo | Loebel et al., 2016 |
| Ris_PT vs Ris | Aman et al., 2009 |
| Memantine vs Ris | Nikvarz et al., 2017 |
| VPA vs Ris | Martsenkovska et al., 2014 |
| Olanz vs Placebo | Hollander et al., 2006 |
| MPH vs Ris | Mahajan et al., 2022 |
| Hal vs Ris | Miral et al., 2008 |
