## Supplementary material 3 for "A network meta-analysis of randomised controlled trials of antipsychotic medications to assess their comparative efficacy and tolerability in autistic people"

1. CYBOCS

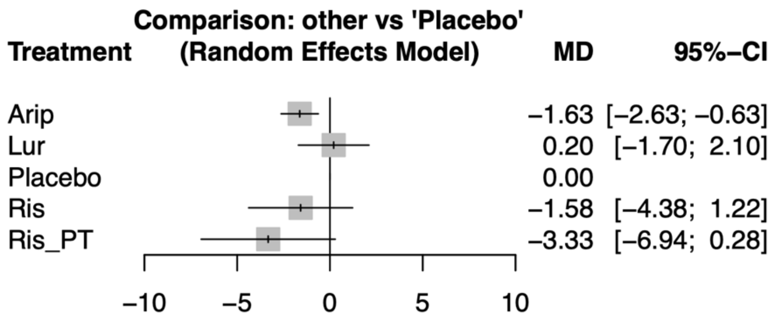


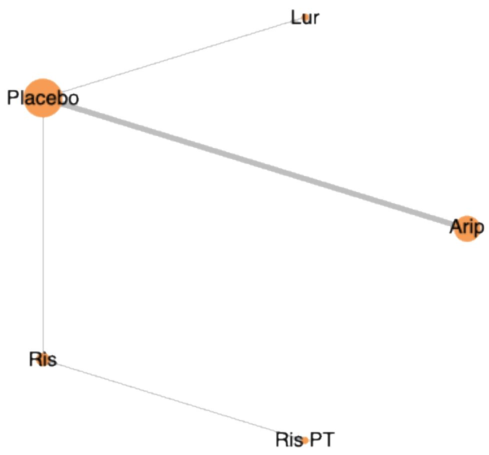


| Rank of interventions | P-scores (random) |
|---|---|
| Ris_PT | 0.9165 |
| Arip | 0.6631 |
| Ris | 0.5669 |
| Placebo | 0.1880 |
| Lur | 0.1655 |

2. Fixed effect NMAs

i. ABC-I

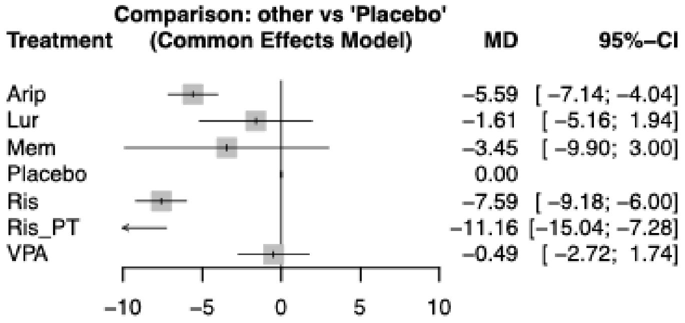


ii. CGI-I

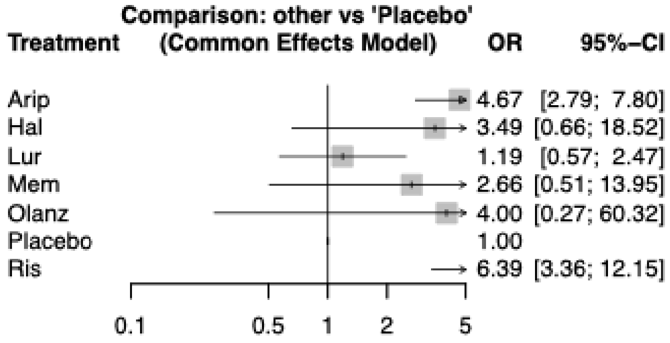


iii. CY-BOCS

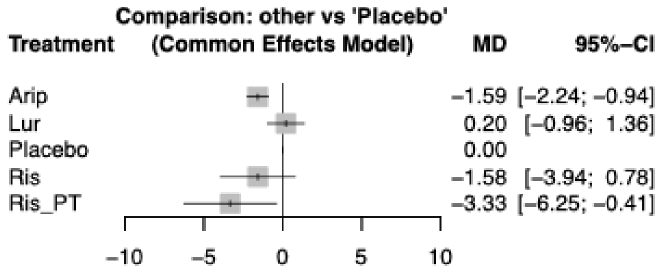


iv. Drop out due to AE

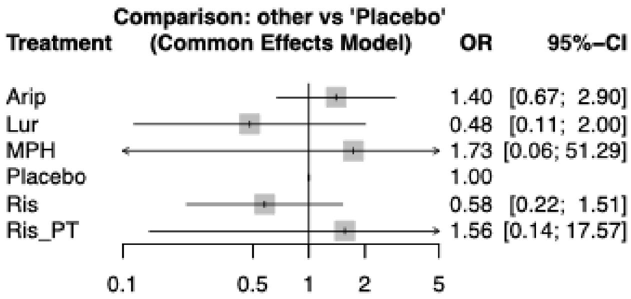

v. Overall AE

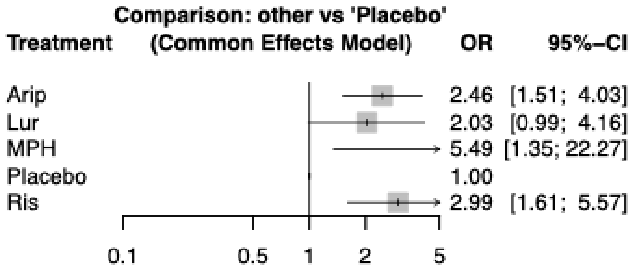


vi. Sedation

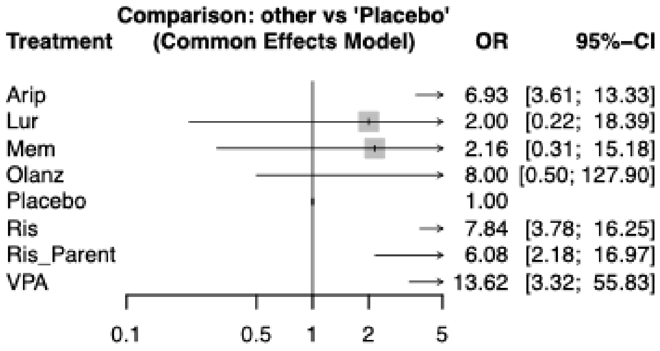


vii. Weight gain

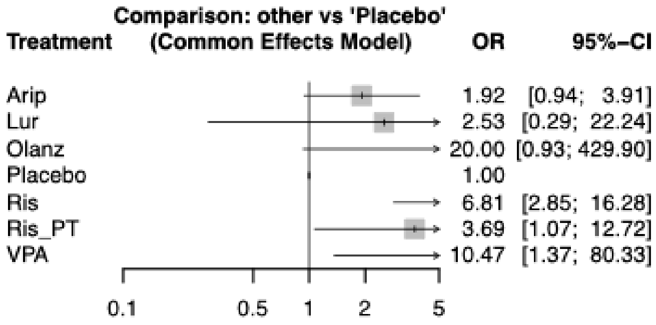

3. CGI-I sensitive analysis

i. Marcus' study removed

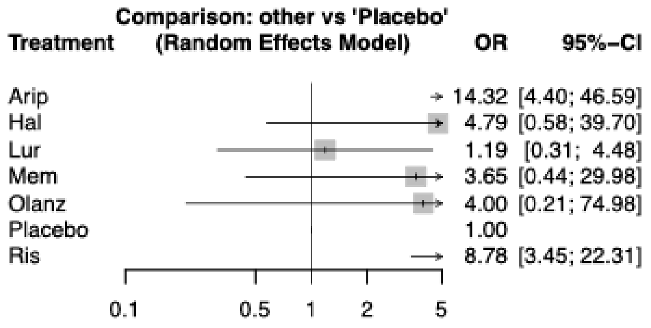


ii. Studies with overall high risk of bias removed (ROB to complete)

| Study name | Number of domains with high risk of bias | Heterogeneity ($I^2$ , $Tau^2$) |
|---|---|---|
| Miral et al., 2008 | 3 | $I^2$ =75.4%, $Tau^2$ =0.9630 |

iii. Studies with the highest impact on the pooled estimates

| Study | Impacts on number of networks (0.99 and above) | Networks |
|---|---|---|
| Nikvarz et al., 2017 | 6 | Arip:Mem; Hal:Mem Lur:Mem; Mem:Olanz; Mem:Placebo; Mem:Ris |
| Loebel et al., 2016 | 6 | Arip:Lur; Lur:Mem; Lur:Olanz; Hal:Lur Lur:Placebo; Lur:Ris |
| Hollander et al., 2006 | 6 | Arip:Olanz; Lur:Olanz; Mem:Olanz; Olanz:Placebo; Olanz:Ris; Hal:Olanz |
| Miral et al., 2008 | 6 | Arip:Hal; Hal:Ris; Hal:Mem; Hal:Lur; Hal:Olanz; Hal:Placebo |

| Networks | Study with highest impact (0.99 and 1.00) |
|---|---|
| Arip:Hal | Miral et al., 2008 |
| Arip:Lur | Loebel et al., 2016 |
| Arip:Mem | Nikvarz et al., 2017 |
| Arip:Olanz | Hollander et al., 2006 |
| Hal:Lur | Loebel et al., 2016 and Miral et al., 2008 |
| Hal:Mem | Nikvarz et al., 2017 and Miral et al., 2008 |
| Hal:Olanz | Hollander et al., 2006 and Miral et al., 2008 |
| Hal:Ris | Miral et al., 2008 |
| Hal:Placebo | Miral et al., 2008 |
| Lur:Olanz | Loebel et al., 2016 and Hollander et al., 2006 |

| | |
|---|---|
| Lur:Placebo | Loebel et al., 2016 |
| Lur:Ris | Loebel et al., 2016 |
| Mem:Olanz | Nikvarz et al., 2017 and Hollander et al., 2006 |
| Mem:Placebo | Nikvarz et al., 2017 |
| Mem:Ris | Nikvarz et al., 2017 |
| Olanz:Placebo | Hollander et al., 2006 |
| Olanz:Ris | Hollander et al., 2006 |

iv. Treatment ranking when individual studies removed

Marcus removed

| Rank of interventions (improvement of symptoms) | P-scores |
|---|---|
| Arip | 0.8796 |
| Ris | 0.7423 |
| Hal | 0.5574 |
| Olanz | 0.5162 |
| Mem | 0.4828 |
| Lur | 0.1941 |
| Placebo | 0.1276 |

Ghanizadeh removed

| Rank of interventions (improvement of symptoms) | P-scores |
|---|---|
| Ris | 0.8236 |
| Hal | 0.6223 |
| Arip | 0.5889 |
| Mem | 0.5553 |
| Olanz | 0.5318 |
| Lur | 0.2305 |
| Placebo | 0.1477 |

Kent removed

| Rank of interventions (improvement of symptoms) | P-scores |
|---|---|
| Ris | 0.7749 |
| Arip | 0.6872 |
| Hal | 0.5866 |
| Mem | 0.5254 |
| Olanz | 0.5146 |
| Lur | 0.2461 |
| Placebo | 0.1652 |

McCracken removed

| Rank of interventions (improvement of symptoms) | P-scores |
|---|---|
| Arip | 0.7690 |
| Ris | 0.7104 |

| | |
|---|---|
| Olanz | 0.5983 |
| Hal | 0.5038 |
| Mem | 0.4313 |
| Lur | 0.2877 |
| Placebo | 0.1994 |

Nikvarz removed

| Rank of interventions (improvement of symptoms) | P-scores |
|---|---|
| Ris | 0.7605 |
| Arip | 0.7275 |
| Hal | 0.5584 |
| Olanz | 0.5506 |
| Lur | 0.2428 |
| Placebo | 0.1602 |

Owen removed

| Rank of interventions (improvement of symptoms) | P-scores |
|---|---|
| Ris | 0.7579 |
| Arip | 0.6715 |
| Hal | 0.5590 |
| Olanz | 0.5573 |
| Mem | 0.4943 |
| Lur | 0.2755 |
| Placebo | 0.1844 |

Loebel removed

| Rank of interventions (improvement of symptoms) | P-scores |
|---|---|
| Ris | 0.7252 |
| Arip | 0.6835 |
| Hal | 0.5180 |
| Olanz | 0.5131 |
| Mem | 0.4459 |
| Placebo | 0.1142 |

Hollander removed

| Rank of interventions (improvement of symptoms) | P-scores |
|---|---|
| Ris | 0.7842 |
| Arip | 0.7440 |
| Hal | 0.5690 |
| Mem | 0.4968 |
| Lur | 0.2460 |
| Placebo | 0.1600 |

McDougle removed

| Rank of interventions (improvement of symptoms) | P-scores |
|---|---|
| Arip | 0.7478 |
| Ris | 0.7305 |
| Olanz | 0.5754 |
| Hal | 0.5282 |
| Mem | 0.4586 |
| Lur | 0.2735 |
| Placebo | 0.1860 |

Miral removed

| Rank of interventions (improvement of symptoms) | P-scores |
|---|---|
| Ris | 0.7752 |
| Arip | 0.7408 |
| Olanz | 0.5604 |
| Mem | 0.5007 |
| Lur | 0.2528 |
| Placebo | 0.1702 |

4. Results of individual studies

i. ABC-I

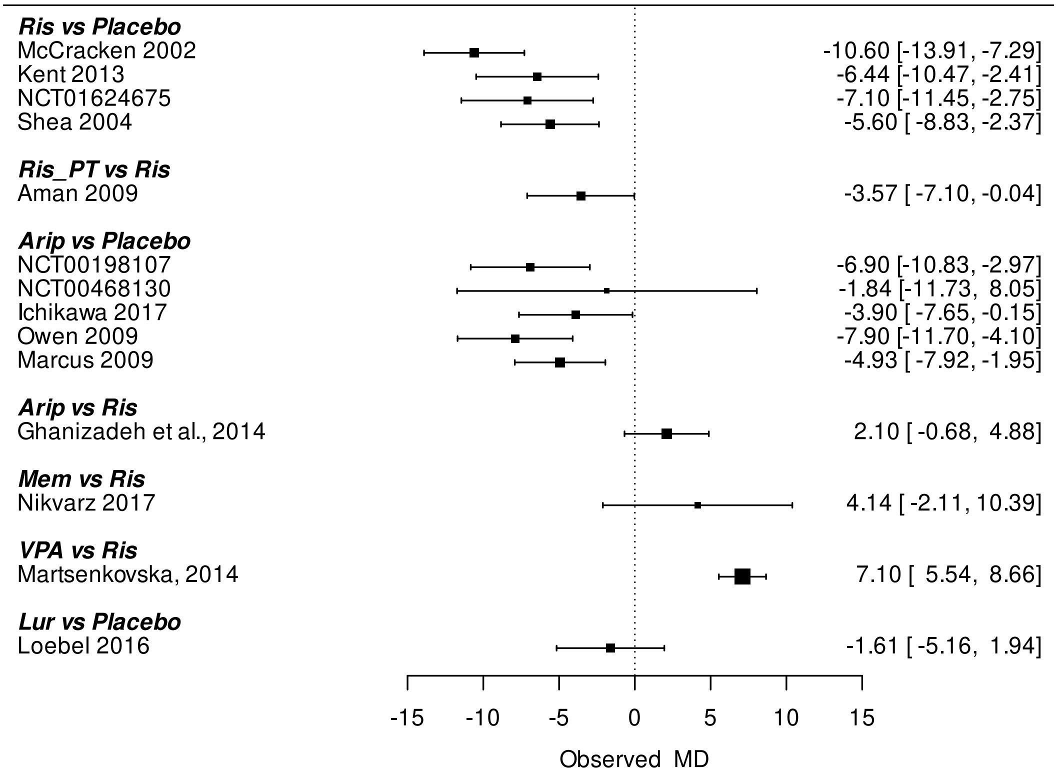


ii. CGI-I

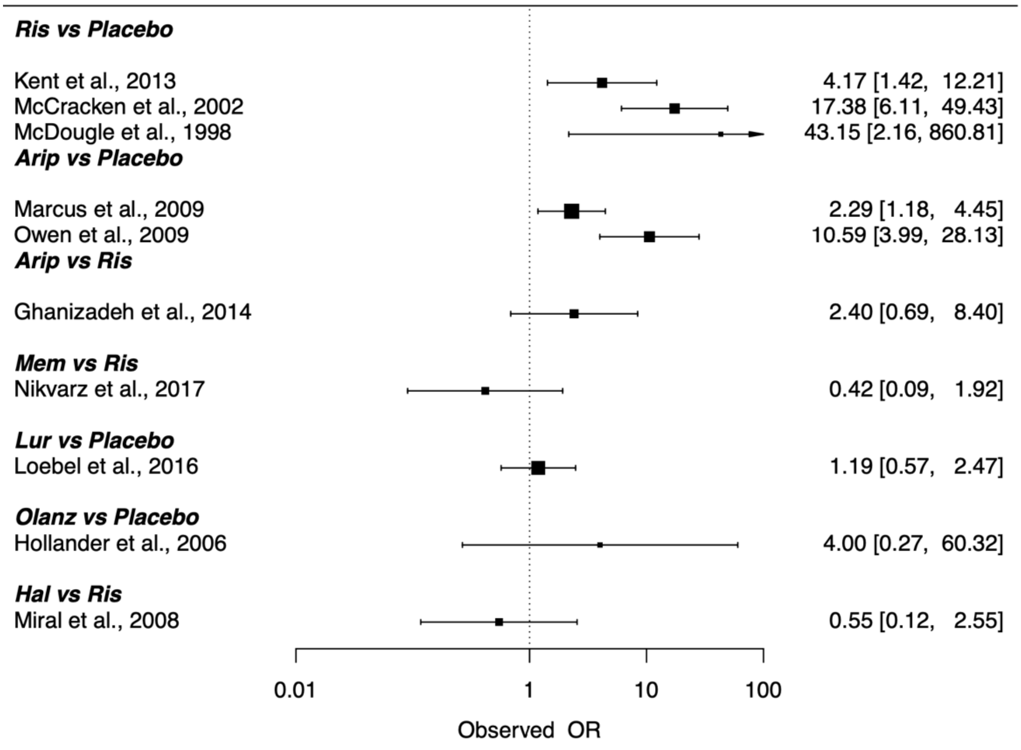

iii. CYBOCS

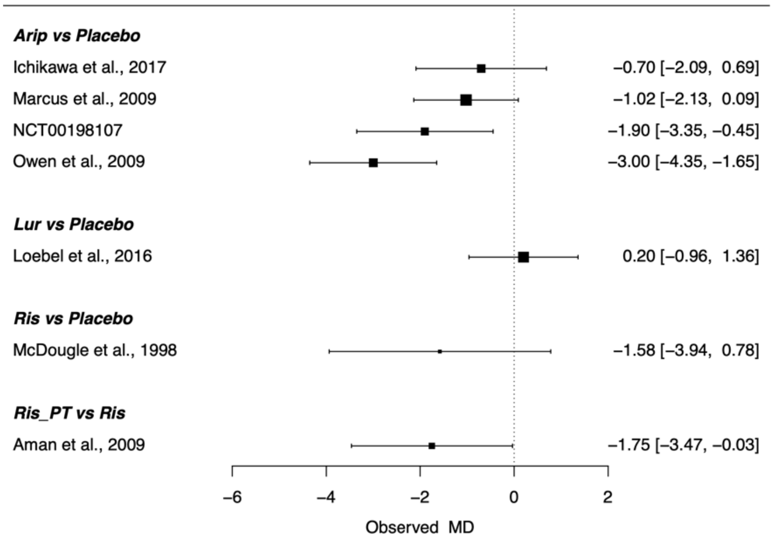


iv. Drop out due to AE

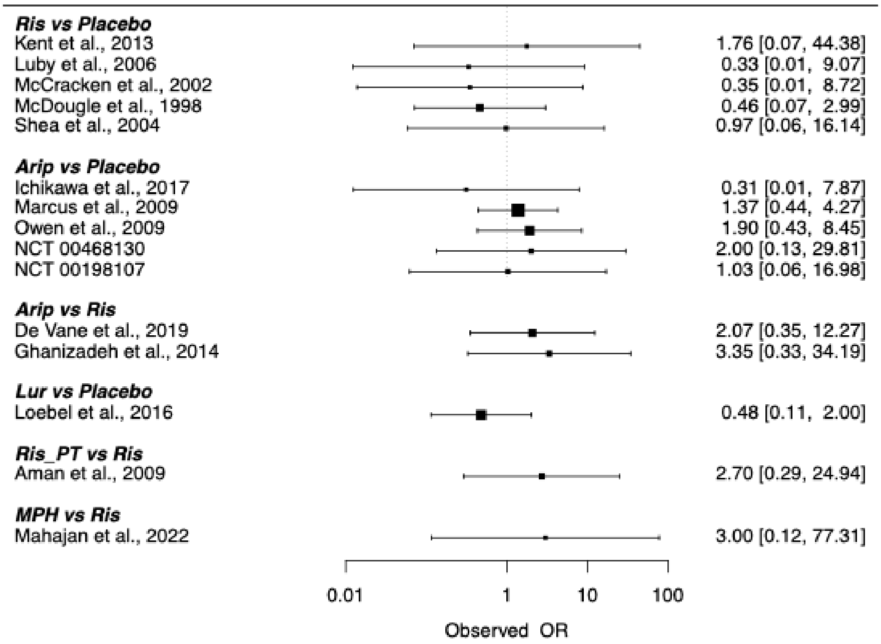


v. Overall AE

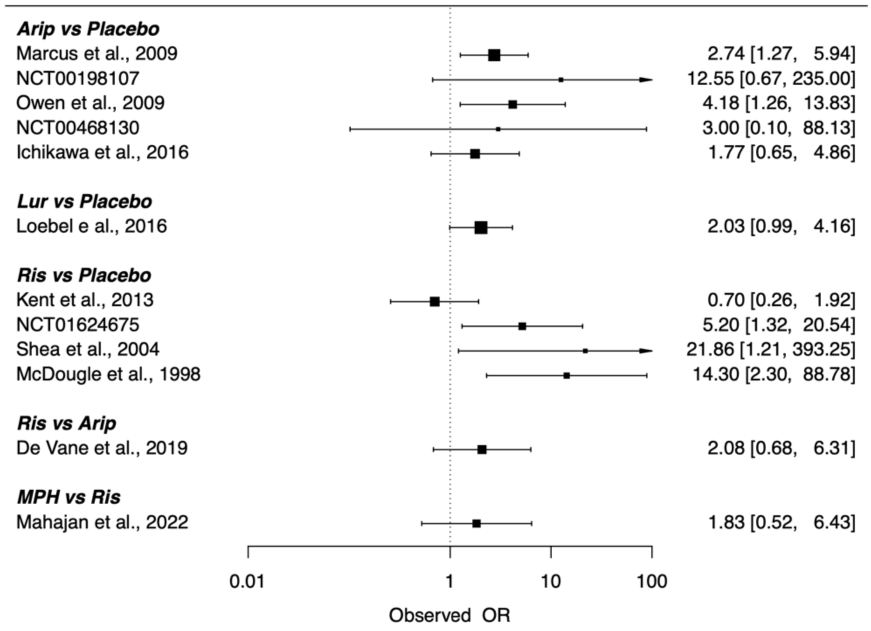


vi. Sedation

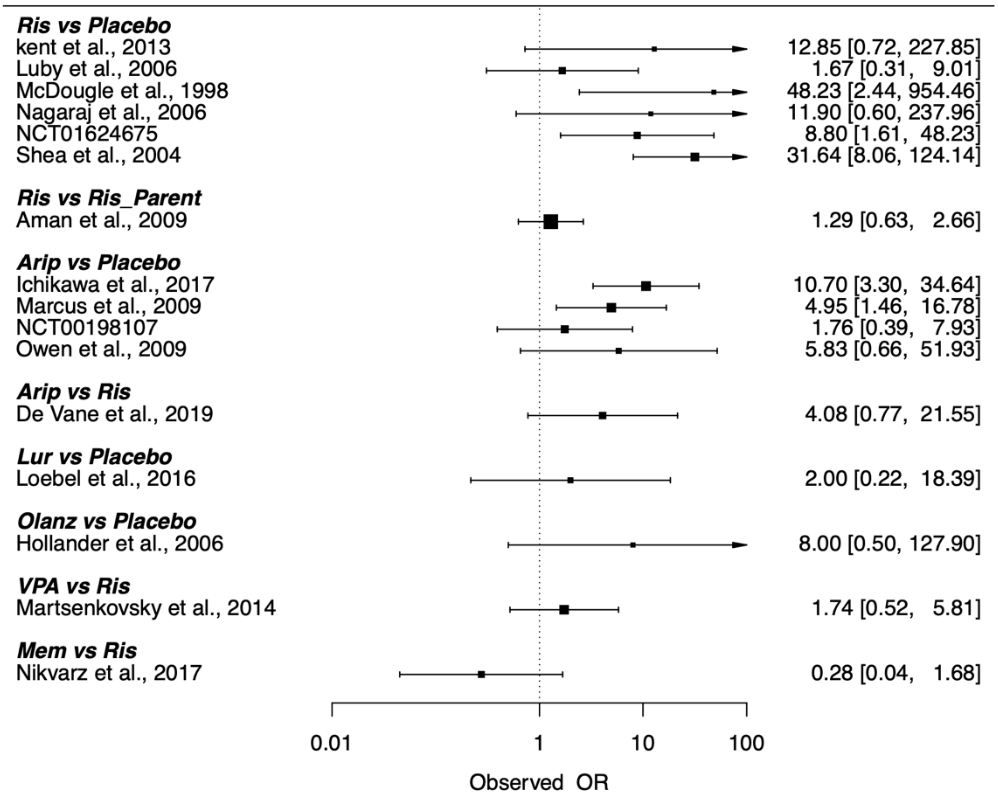


vii. Weight gain

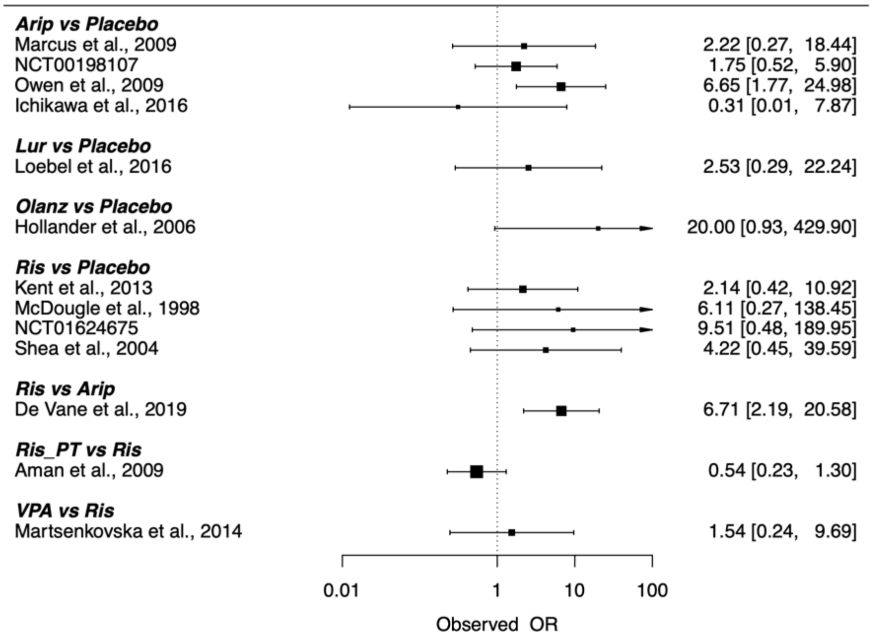
Arip vs Placebo
Marcus et al., 2009
2.22 [0.27, 18.44]
NCT00198107
1.75 [0.52, 5.90]
Owen et al., 2009
6.65 [1.77, 24.98]
Ichikawa et al., 2016
0.31 [0.01, 7.87]
Lur vs Placebo
Loebel et al., 2016
2.53 [0.29, 22.24]
Olanz vs Placebo
Hollander et al., 2006
20.00 [0.93, 429.90]
Ris vs Placebo
Kent et al., 2013
2.14 [0.42, 10.92]
McDougle et al., 1998
6.11 [0.27, 138.45]
NCT01624675
9.51 [0.48, 189.95]
Shea et al., 2004
4.22 [0.45, 39.59]
Ris vs Arip
De Vane et al., 2019
6.71 [2.19, 20.58]
Ris_PT vs Ris
Aman et al., 2009
0.54 [0.23, 1.30]
VPA vs Ris
Martsenkovska et al., 2014
1.54 [0.24, 9.69]
0.01
1
10
100
Observed OR

5. Inconsistency results

### ABC-I

| | Comparison | No.Studies | NMA | Direct | Indirect | Difference | Diff_95CI_lower | Diff_95CI_upper | pValue |
|---|---|---|---|---|---|---|---|---|---|
| 1 | Arip:Lur | 0 | -3.98166125116952 | NA | -3.98166125116952 | NA | NA | NA | NA |
| 2 | Arip:Mem | 0 | -2.1441238124881 | NA | -2.1441238124881 | NA | NA | NA | NA |
| 3 | Arip:Placebo | 5 | -5.59166125116952 | -5.63156552190098 | -5.44420512925927 | -0.187360392641709 | -4.06958836512933 | 3.69486757984592 | 0.924640558990128 |
| 4 | Arip:Ris | 1 | 1.99587618751191 | 2.1 | 1.91263960735827 | 0.187360392641726 | -3.69486757984459 | 4.06958836512935 | 0.924640558990121 |
| 5 | Arip:Ris_PT | 0 | 5.5658761875119 | NA | 5.5658761875119 | NA | NA | NA | NA |
| 6 | Arip:VPA | 0 | -5.10412381248809 | NA | -5.10412381248809 | NA | NA | NA | NA |
| 7 | Lur:Mem | 0 | 1.83753743868142 | NA | 1.83753743868142 | NA | NA | NA | NA |
| 8 | Lur:Placebo | 1 | -1.61 | -1.61 | NA | NA | NA | NA | NA |
| 9 | Lur:Ris | 0 | 5.97753743868143 | NA | 5.97753743868143 | NA | NA | NA | NA |
| 10 | Lur:Ris_PT | 0 | 9.54753743868143 | NA | 9.54753743868143 | NA | NA | NA | NA |
| 11 | Lur:VPA | 0 | -1.12246256131857 | NA | -1.12246256131857 | NA | NA | NA | NA |
| 12 | Mem:Placebo | 0 | -3.44753743868143 | NA | -3.44753743868143 | NA | NA | NA | NA |
| 13 | Mem:Ris | 1 | 4.14000000000001 | 4.14 | NA | NA | NA | NA | NA |
| 14 | Mem:Ris_PT | 0 | 7.71 | NA | 7.71 | NA | NA | NA | NA |
| 15 | Mem:VPA | 0 | -2.95999999999999 | NA | -2.95999999999999 | NA | NA | NA | NA |
| 16 | Ris:Placebo | 4 | -7.58753743868143 | -7.54420512925925 | -7.73156552190097 | 0.187360392641722 | -3.69486757984459 | 4.06958836512935 | 0.924640558990123 |
| 17 | Ris_PT:Placebo | 0 | -11.1575374386814 | NA | -11.1575374386814 | NA | NA | NA | NA |
| 18 | VPA:Placebo | 0 | -0.487537438681432 | NA | -0.487537438681432 | NA | NA | NA | NA |
| 19 | Ris:Ris_PT | 1 | 3.56999999999999 | 3.57 | NA | NA | NA | NA | NA |

| | | | | | | | | | |
|---|---|---|---|---|---|---|---|---|---|
| 20 | Ris:VPA | 1 | -7.1 | -7.1 | NA | NA | NA | NA | NA |
| 21 | Ris_PT:VPA | 0 | -10.67 | NA | -10.67 | NA | NA | NA | NA |

### CGI-I

| | Comparison | No.Studies | NMA | Direct | Indirect | Difference | Diff_95CI_lower | Diff_95CI_upper | pValue |
|---|---|---|---|---|---|---|---|---|---|
| 1 | Arip:Hal | 0 | 0.520181633899999 | NA | 0.520181633899999 | NA | NA | NA | NA |
| 2 | Arip:Lur | 0 | 1.7790074221142 | NA | 1.7790074221142 | NA | NA | NA | NA |
| 3 | Arip:Mem | 0 | 0.792115349383642 | NA | 0.792115349383642 | NA | NA | NA | NA |
| 4 | Arip:Olanz | 0 | 0.562996639228787 | NA | 0.562996639228787 | NA | NA | NA | NA |
| 5 | Arip:Placebo | 2 | 1.94929100034868 | 1.54945927802992 | 3.28045314171519 | -1.73099386368527 | -4.81070392415809 | 1.34871619678755 | 0.270624730113478 |
| 6 | Arip:Ris | 1 | -0.0859541696703152 | 0.8754687373539 | -0.855525126331371 | 1.73099386368527 | -1.34871619678755 | 4.81070392415809 | 0.270624730113478 |
| 7 | Hal:Lur | 0 | 1.2588257882142 | NA | 1.2588257882142 | NA | NA | NA | NA |
| 8 | Hal:Mem | 0 | 0.271933715483643 | NA | 0.271933715483643 | NA | NA | NA | NA |
| 9 | Hal:Olanz | 0 | 0.0428150053287881 | NA | 0.0428150053287881 | NA | NA | NA | NA |
| 10 | Hal:Placebo | 0 | 1.42910936644868 | NA | 1.42910936644868 | NA | NA | NA | NA |
| 11 | Hal:Ris | 1 | -0.606135803570314 | -0.606135803570316 | NA | NA | NA | NA | NA |
| 12 | Lur:Mem | 0 | -0.986892072730559 | NA | -0.986892072730559 | NA | NA | NA | NA |
| 13 | Lur:Olanz | 0 | -1.21601078288541 | NA | -1.21601078288541 | NA | NA | NA | NA |
| 14 | Lur:Placebo | 1 | 0.170283578234475 | 0.170283578234476 | NA | NA | NA | NA | NA |
| 15 | Lur:Ris | 0 | -1.86496159178452 | NA | -1.86496159178452 | NA | NA | NA | NA |
| 16 | Mem:Olanz | 0 | -0.229118710154855 | NA | -0.229118710154855 | NA | NA | NA | NA |
| 17 | Mem:Placebo | 0 | 1.15717565096503 | NA | 1.15717565096503 | NA | NA | NA | NA |
| 18 | Mem:Ris | 1 | -0.878069519053957 | -0.878069519053957 | NA | NA | NA | NA | NA |

| | | | | | | | | | |
|---|---|---|---|---|---|---|---|---|---|
| 19 | Olanz:Placebo | 1 | 1.38629436111989 | 1.38629436111989 | NA | NA | NA | NA | NA |
| 20 | Olanz:Ris | 0 | -0.648950808899103 | NA | -0.648950808899103 | NA | NA | NA | NA |
| 21 | Ris:Placebo | 3 | 2.03524517001899 | 2.40498440436129 | 0.673990540676019 | 1.73099386368527 | -1.34871619678755 | 4.81070392415809 | 0.270624730113478 |

### CYBOCS

| | Comparison | No.Studies | NMA | Direct | Indirect | Difference | Diff_95CI_lower | Diff_95CI_upper | pValue |
|---|---|---|---|---|---|---|---|---|---|
| 1 | Arip:Lur | 0 | -1.83090636487699 | NA | -1.83090636487699 | NA | NA | NA | NA |
| 2 | Arip:Placebo | 4 | -1.63090636487699 | -1.63090636487699 | NA | NA | NA | NA | NA |
| 3 | Arip:Ris | 0 | -0.0509063648769883 | NA | -0.0509063648769883 | NA | NA | NA | NA |
| 4 | Arip:Ris_PT | 0 | 1.69909363512301 | NA | 1.69909363512301 | NA | NA | NA | NA |
| 5 | Lur:Placebo | 1 | 0.2 | 0.2 | NA | NA | NA | NA | NA |
| 6 | Lur:Ris | 0 | 1.78 | NA | 1.78 | NA | NA | NA | NA |
| 7 | Lur:Ris_PT | 0 | 3.53 | NA | 3.53 | NA | NA | NA | NA |
| 8 | Ris:Placebo | 1 | -1.58 | -1.58 | NA | NA | NA | NA | NA |
| 9 | Ris_PT:Placebo | 0 | -3.33 | NA | -3.33 | NA | NA | NA | NA |
| 10 | Ris:Ris_PT | 1 | 1.75 | 1.75 | NA | NA | NA | NA | NA |

### Drop out due to AE

| | Comparison | No.Studies | NMA | Direct | Indirect | Difference | Diff_95CI_lower | Diff_95CI_upper | pValue |
|---|---|---|---|---|---|---|---|---|---|
| 1 | Arip:Lur | 0 | 1.07046480829539 | NA | 1.07046480829539 | NA | NA | NA | NA |
| 2 | Arip:MPH | 0 | -0.213713090922992 | NA | -0.213713090922992 | NA | NA | NA | NA |
| 3 | Arip:Placebo | 5 | 0.334758013316653 | 0.328060304759296 | 0.371147519918083 | -0.043087215158787 | -2.05816269701282 | 1.97198826669524 | 0.966571403332805 |
| 4 | Arip:Ris | 2 | 0.884899197745118 | 0.906057748512576 | 0.862970533353791 | 0.0430872151587853 | -1.97198826669525 | 2.05816269701282 | 0.966571403332806 |
| 5 | Arip:Ris_PT | 0 | -0.109916297241348 | NA | -0.109916297241348 | NA | NA | NA | NA |
| 6 | Lur:MPH | 0 | -1.28417789921839 | NA | -1.28417789921839 | NA | NA | NA | NA |

| | | | | | | | | | |
|---|---|---|---|---|---|---|---|---|---|
| 7 | Lur:Placebo | 1 | -0.735706794978741 | -0.735706794978741 | NA | NA | NA | NA | NA |
| 8 | Lur:Ris | 0 | -0.185565610550276 | NA | -0.185565610550276 | NA | NA | NA | NA |
| 9 | Lur:Ris_PT | 0 | -1.18038110553674 | NA | -1.18038110553674 | NA | NA | NA | NA |
| 10 | MPH:Placebo | 0 | 0.548471104239645 | NA | 0.548471104239645 | NA | NA | NA | NA |
| 11 | MPH:Ris | 1 | 1.09861228866811 | 1.09861228866811 | NA | NA | NA | NA | NA |
| 12 | MPH:Ris_PT | 0 | 0.103796793681644 | NA | 0.103796793681644 | NA | NA | NA | NA |
| 13 | Ris:Placebo | 5 | -0.550141184428465 | -0.534910228594494 | -0.57799744375328 | 0.0430872151587857 | -1.97198826669525 | 2.05816269701282 | 0.966571403332806 |
| 14 | Ris_PT:Placebo | 0 | 0.444674310558001 | NA | 0.444674310558001 | NA | NA | NA | NA |
| 15 | Ris:Ris_PT | 1 | -0.994815494986466 | -0.994815494986466 | NA | NA | NA | NA | NA |

### Overall AE

| | Comparison | No.Studies | NMA | Direct | Indirect | Difference | Diff_95CI_lower | Diff_95CI_upper | pValue |
|---|---|---|---|---|---|---|---|---|---|
| 1 | Arip:Lur | 0 | 0.313363009113345 | NA | 0.313363009113345 | NA | NA | NA | NA |
| 2 | Arip:MPH | 0 | -0.971930496659668 | NA | -0.971930496659668 | NA | NA | NA | NA |
| 3 | Arip:Placebo | 5 | 1.02030351180563 | 1.1158923607996 | 0.526739509452049 | 0.589152851347549 | -1.67432511833726 | 2.85263082103236 | 0.609944804854079 |
| 4 | Arip:Ris | 1 | -0.365794693089352 | -0.730051737495396 | -0.140898886147844 | -0.589152851347552 | -2.85263082103236 | 1.67432511833726 | 0.609944804854077 |
| 5 | Lur:MPH | 0 | -1.28529350577301 | NA | -1.28529350577301 | NA | NA | NA | NA |
| 6 | Lur:Placebo | 1 | 0.706940502692282 | 0.706940502692281 | NA | NA | NA | NA | NA |
| 7 | Lur:Ris | 0 | -0.679157702202697 | NA | -0.679157702202697 | NA | NA | NA | NA |
| 8 | MPH:Placebo | 0 | 1.99223400846529 | NA | 1.99223400846529 | NA | NA | NA | NA |
| 9 | MPH:Ris | 1 | 0.606135803570316 | 0.606135803570315 | NA | NA | NA | NA | NA |
| 10 | Ris:Placebo | 4 | 1.38609820489498 | 1.25679124694744 | 1.84594409829499 | -0.589152851347549 | -2.85263082103236 | 1.67432511833726 | 0.609944804854079 |

### Sedation

| | Comparison | No.Studies | NMA | Direct | Indirect | Difference | Diff_95CI_lower | Diff_95CI_upper | pValue |
|---|---|---|---|---|---|---|---|---|---|
| 1 | Arip:Lur | 0 | 1.24923317839335 | NA | 1.24923317839335 | NA | NA | NA | NA |
| 2 | Arip:Mem | 0 | 1.17264717623436 | NA | 1.17264717623436 | NA | NA | NA | NA |
| 3 | Arip:Olanz | 0 | -0.137061182726547 | NA | -0.137061182726547 | NA | NA | NA | NA |
| 4 | Arip:Placebo | 4 | 1.94238035895329 | 1.60598045657427 | 3.80218278728079 | -2.19620233070652 | -4.90592800562238 | 0.513523344209331 | 0.112166797581422 |
| 5 | Arip:Ris | 1 | -0.118337005081206 | 1.40691364832263 | -0.789288682383901 | 2.19620233070653 | -0.513523344209328 | 4.90592800562238 | 0.112166797581422 |
| 6 | Arip:Ris_Parent | 0 | 0.136305213292375 | NA | 0.136305213292375 | NA | NA | NA | NA |
| 7 | Arip:VPA | 0 | -0.670578732563913 | NA | -0.670578732563913 | NA | NA | NA | NA |
| 8 | Lur:Mem | 0 | -0.0765860021589844 | NA | -0.0765860021589844 | NA | NA | NA | NA |
| 9 | Lur:Olanz | 0 | -1.38629436111989 | NA | -1.38629436111989 | NA | NA | NA | NA |
| 10 | Lur:Placebo | 1 | 0.693147180559947 | 0.693147180559945 | NA | NA | NA | NA | NA |
| 11 | Lur:Ris | 0 | -1.36757018347455 | NA | -1.36757018347455 | NA | NA | NA | NA |
| 12 | Lur:Ris_Parent | 0 | -1.11292796510097 | NA | -1.11292796510097 | NA | NA | NA | NA |
| 13 | Lur:VPA | 0 | -1.91981191095726 | NA | -1.91981191095726 | NA | NA | NA | NA |
| 14 | Mem:Olanz | 0 | -1.30970835896091 | NA | -1.30970835896091 | NA | NA | NA | NA |
| 15 | Mem:Placebo | 0 | 0.769733182718931 | NA | 0.769733182718931 | NA | NA | NA | NA |
| 16 | Mem:Ris | 1 | -1.29098418131557 | -1.29098418131557 | NA | NA | NA | NA | NA |
| 17 | Mem:Ris_Parent | 0 | -1.03634196294199 | NA | -1.03634196294199 | NA | NA | NA | NA |
| 18 | Mem:VPA | 0 | -1.84322590879827 | NA | -1.84322590879827 | NA | NA | NA | NA |

| | | | | | | | | | |
|---|---|---|---|---|---|---|---|---|---|
| 19 | Olanz:Placebo | 1 | 2.07944154167984 | 2.07944154167984 | NA | NA | NA | NA | NA |
| 20 | Olanz:Ris | 0 | 0.0187241776453413 | NA | 0.0187241776453413 | NA | NA | NA | NA |
| 21 | Olanz:Ris_Parent | 0 | 0.273366396018922 | NA | 0.273366396018922 | NA | NA | NA | NA |
| 22 | Olanz:VPA | 0 | -0.533517549837366 | NA | -0.533517549837366 | NA | NA | NA | NA |
| 23 | Ris:Placebo | 6 | 2.0607173640345 | 2.39526913895817 | 0.199066808251638 | 2.19620233070653 | -0.513523344209323 | 4.90592800562239 | 0.112166797581421 |
| 24 | Ris_Parent:Placebo | 0 | 1.80607514566092 | NA | 1.80607514566092 | NA | NA | NA | NA |
| 25 | VPA:Placebo | 0 | 2.61295909151721 | NA | 2.61295909151721 | NA | NA | NA | NA |
| 26 | Ris:Ris_Parent | 1 | 0.254642218373581 | 0.254642218373581 | NA | NA | NA | NA | NA |
| 27 | Ris:VPA | 1 | -0.552241727482708 | -0.552241727482708 | NA | NA | NA | NA | NA |
| 28 | Ris_Parent:VPA | 0 | -0.806883945856288 | NA | -0.806883945856288 | NA | NA | NA | NA |

### Weight gain

| | Comparison | No.Studies | NMA | Direct | Indirect | Difference | Diff_95CI_lower | Diff_95CI_upper | pValue |
|---|---|---|---|---|---|---|---|---|---|
| 1 | Arip:Lur | 0 | -0.291048708020452 | NA | -0.291048708020452 | NA | NA | NA | NA |
| 2 | Arip:Olanz | 0 | -2.36001894983299 | NA | -2.36001894983299 | NA | NA | NA | NA |
| 3 | Arip:Placebo | 4 | 0.635713323721 | 0.947565929254067 | -0.596183445794254 | 1.54374937504832 | -0.453191894606339 | 3.54069064470298 | 0.129730921165918 |
| 4 | Arip:Ris | 1 | -1.22321326176778 | -1.90335053463652 | -0.359601159588199 | -1.54374937504832 | -3.54069064470298 | 0.453191894606342 | 0.129730921165919 |
| 5 | Arip:Ris_PT | 0 | -0.610807845183867 | NA | -0.610807845183867 | NA | NA | NA | NA |
| 6 | Arip:VPA | 0 | -1.65337098246631 | NA | -1.65337098246631 | NA | NA | NA | NA |
| 7 | Lur:Olanz | 0 | -2.06897024181254 | NA | -2.06897024181254 | NA | NA | NA | NA |
| 8 | Lur:Placebo | 1 | 0.926762031741452 | 0.92676203174145 | NA | NA | NA | NA | NA |
| 9 | Lur:Ris | 0 | -0.932164553747326 | NA | -0.932164553747326 | NA | NA | NA | NA |

| | | | | | | | | | |
|---|---|---|---|---|---|---|---|---|---|
| 10 | Lur:Ris_PT | 0 | -0.319759137163415 | NA | -0.319759137163415 | NA | NA | NA | NA |
| 11 | Lur:VPA | 0 | -1.36232227444586 | NA | -1.36232227444586 | NA | NA | NA | NA |
| 12 | Olanz:Placebo | 1 | 2.99573227355399 | 2.99573227355399 | NA | NA | NA | NA | NA |
| 13 | Olanz:Ris | 0 | 1.13680568806521 | NA | 1.13680568806521 | NA | NA | NA | NA |
| 14 | Olanz:Ris_PT | 0 | 1.74921110464913 | NA | 1.74921110464913 | NA | NA | NA | NA |
| 15 | Olanz:VPA | 0 | 0.706647967366679 | NA | 0.706647967366679 | NA | NA | NA | NA |
| 16 | Ris:Placebo | 4 | 1.85892658548878 | 1.30716708884226 | 2.85091646389059 | -1.54374937504832 | -3.54069064470298 | 0.453191894606338 | 0.129730921165918 |
| 17 | Ris_PT:Placebo | 0 | 1.24652116890487 | NA | 1.24652116890487 | NA | NA | NA | NA |
| 18 | VPA:Placebo | 0 | 2.28908430618731 | NA | 2.28908430618731 | NA | NA | NA | NA |
| 19 | Ris:Ris_PT | 1 | 0.612405416583911 | 0.612405416583911 | NA | NA | NA | NA | NA |
| 20 | Ris:VPA | 1 | -0.430157720698537 | -0.430157720698536 | NA | NA | NA | NA | NA |
| 21 | Ris_PT:VPA | 0 | -1.04256313728245 | NA | -1.04256313728245 | NA | NA | NA | NA |
